# iSCORED: nanopore-based random genomic sampling for intraoperative molecular diagnosis

**DOI:** 10.1101/2023.10.17.23297170

**Authors:** Francesco E. Emiliani, Abdol Aziz Ould Ismail, Edward G. Hughes, Gregory J. Tsongalis, George J Zanazzi, Chun-Chieh Lin

## Abstract

Copy number variations (CNVs) are almost ubiquitous in cancer. In many cases, somatic CNV analysis has led to the identification of oncogenic pathways and suggested molecular-defined therapeutic targets. Here, we develop iSCORED, a one-step random genomic DNA reconstruction method that enables efficient, unbiased quantification of CNVs using a real-time Nanopore sequencer. By leveraging the long concatenated reads, we generate approximately 1-2 million genomic fragments within one hour of MinION sequencing, allowing for high-resolution genomic dosage comparisons. In our cohort of 26 malignant brain tumors, we demonstrated 100% concordance in CNV detections, including chromosomal alterations and oncogene amplifications when compared to clinically validated next generation sequencing and chromosomal microarray results. In addition, iSCORED allows concurrent brain tumor methylation classification without additional tissue preparation. The integrated methylation information revealed promoter hypomethylation in all detected amplified oncogenes. The entire workflow, including the automatic generation of CNV and methylation reports, can be accomplished within 120-140 minutes. Ultrafast molecular analysis can enhance clinical decision-making, optimize surgical planning and identify potential molecular therapies within surgical timeframes.

## Background

Copy number variations (CNVs) contribute to cancer development and progression by activating oncogenes and inactivating tumor suppressor genes^1–3^. As a predominant class of genomic alterations, CNVs are also involved in a wide range of biological processes, including human evolution^4–6^, neurodegeneration^7,8^, and developmental disorders^4,9–12^. Despite the importance of CNVs in genomic biology, the existing CNV analysis relies on nucleotide hybridization^5–7,10,11,13–15^ and next-generation sequencing^1–3,8^, necessitating high-complexity centralized laboratories with a turnaround time of several days to weeks. Besides the high costs, the prolonged process could delay clinical molecular diagnosis and therapeutic plans^16,17^. In contrast, Nanopore sequencing (Oxford Nanopore Technologies, ONT) is an inexpensive and portable device that provides real-time interpretation of long-read nucleotide sequences. While there have been some successes in ultrafast CNV diagnostics^18,19^, the genomic resolution of short-read sequencing (STORK^19^) is restricted to 10Mb. This limitation is primarily due to low numbers of aligned DNA fragments obtainable within a short sequencing timeframe.

Ultrafast high-resolution CNV detection can be achieved by analyzing randomly concatenated DNA fragments. The approach enables the identification of multiple mappable DNA fragments in one sequencing read, thus optimizing sequencing efficacy. By sequencing a fraction of randomly assembled genomic fragments, the genome-wide chromosomal integrity can be quantitatively assessed. While previous attempts using long concatenated reads for quantitative genomic analysis have been introduced, the methods are lengthy and require sequential mechanical shearing (SMASH^20^) or enzymatic digestion (SMURF ^21,22^) followed by DNA purification, and ligation. The multi-step preparation requires high amount of input DNA and takes a few hours for processing. An important rate-limiting factor has been the lack of a highly efficient method to reliably process genomic DNA *in one reaction*.

Here we develop a novel one-step random genomic DNA concatenation method, named irreversible Sticking Compatible Overhang to Reconstruct DNA (iSCORED), which enables efficient and unbiased CNV assessment using a real-time Nanopore sequencer. This is achieved by analyzing the long concatenated reads (∼1-2 kb) assembled from short DNA fragments (∼100-150 bp), thus augmenting the quantity of mappable DNA fragments per sequencing read. Furthermore, by utilizing the 5-methylcytosine data available through Nanopore sequencing, we demonstrated the feasibility of concurrent methylation classification in primary CNS tumors and promoter methylation analysis of amplified oncogenes. The pipeline was applied to a cohort of 26 intracranial neoplasms, consisting of 17 primary CNS tumors and 9 metastatic tumors. The results were compared to those from next-generation sequencing (TruSight® Tumor 170 and whole exome sequencing) and chromosomal microarray (Affymetrix OncoScan®)^23^ generated in a Clinical Laboratory Improvement Amendments (CLIA) certified pathology laboratory at Dartmouth-Hitchcock Medical Center.

## Results

### Concurrent fragmentation and concatenation of genomic DNA

The central concept of iSCORED is simultaneous digestion and ligation of DNA molecules by utilizing a panel of restriction endonucleases (REs) capable of generating compatible cohesive ends. Within the same reaction, DNA ligase catalyzes random re-ligation of the digested fragments to form long concatemers. Irreversible ligation products are generated when cohesive ends produced by different restriction enzymes are ligated together. This unidirectional reaction is possible because of the staggered nature of the DNA recognition sequences and actual phosphodiester bond breakage sites (Fig. 1a, b, Figure S1). The likelihood of forming such irreversible ligations increases with the number of different restriction enzymes producing compatible ends. Using CTAG overhangs as an example, the digested fragments were concatenated to larger chimeric molecules in the presence of DNA ligase (Figure S1).

**Fig. 1:**
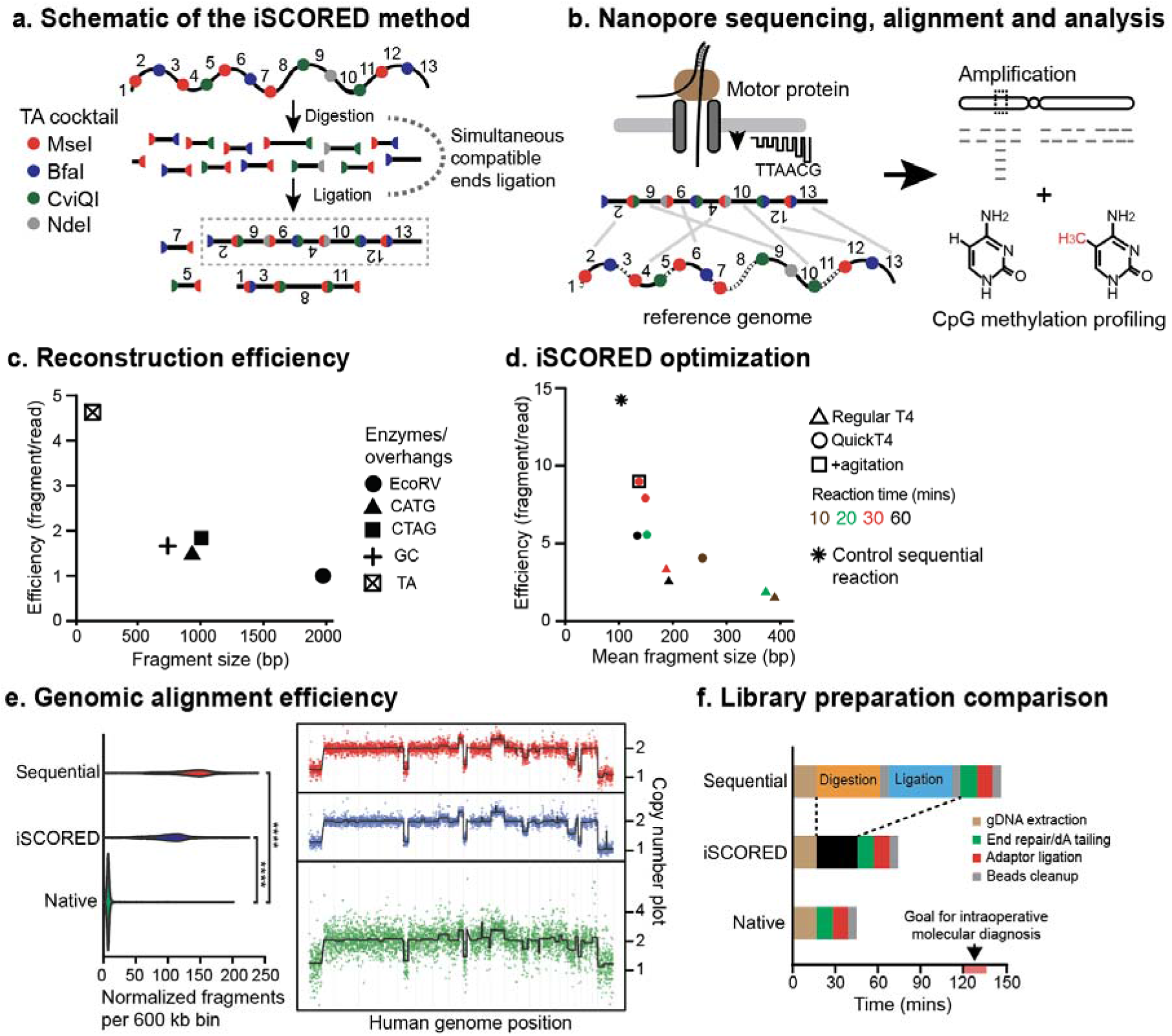
Proposed iSCORED method for ultrafast copy number analysis. **a**, iSCORED schematic showing simultaneous compatible end ligation with TA enzyme cocktail (T^TAA, C^TAG, G^TAC and CA^TATG by MseI, BfaI, CviQI and NdeI, respectively). **b**, Long stochastically concatenated DNA molecules are analyzed with Nanopore device and aligned to the reference for genome-wide quantitative measurement. **c**, The reconstruction efficiency of four iSCORED cocktail combinations are compared. The reaction is incubated at 37 °C for 30 mins. CATG cocktail: NcoI (C^CATGG), PciI (A^CATGT), BspHI (T^CATGA). CTAG cocktail: NheI (G^CTAGC), SpeI (A^CTAGT), AvrII (C^CTAGG), XbaI (T^CTAGA). CG cocktail: MspI (C^CGG), HinP1I (G^CGC), HpyCH4IV (A^CGT) and TaqI-V2 (T^CGA). EcoRV is employed as a control since it generates blunt ends upon restriction digestion. **d**, Optimization of iSCORED reaction by adjusting various experimental parameters, such as incubation periods, DNA ligases and intermittent mixing and cooling. **e**, An oligodendroglioma sample was processed either sequentially (digestion, purification, and ligation), with iSCORED, or sequenced as native gDNA. Samples were normalized to contain the same amount of sequencing data. The number of unique fragments mapped per genomic bin are shown for each sample (left panels). The resulting CNV plots are shown in the right panels (resolution=600kb per bin). CoV for sequential approach, iSCORED and native gDNA sequencing are 0.57, 0.54 and 3.3, respectively. **f**, Comparison of library preparation times across three methods. The sequential DNA digestion and ligation required 150 minutes, while the iSCORED required 75 minutes and the native DNA method required 45 minutes. The goal of intraoperative molecular diagnosis is achieved within 120-150 minutes of receiving the resected specimen.

### Systematic analysis and optimization of all overhang candidates

We next examined all existing 4-mer and 6-mer Type IIP REs capable of generating 2-nucleotide and 4-nucleotide overhangs (Figure S2). Given the palindromic nature of Type IIP REs, there are 16 (=4^2^) and 4 (=4^1^) possible combinations for 4-nucleotide and 2-nucleotide overhangs, respectively. Depending on the RE recognition sequence (4 or 6 bp), the same overhang could be generated by 4^1^ or 4^2^ different enzymes; however, some of the theoretical combinations do not exist, and some are partially or completely blocked by DNA methylation (Figure 2; New England Biolabs).

**Fig. 2:**
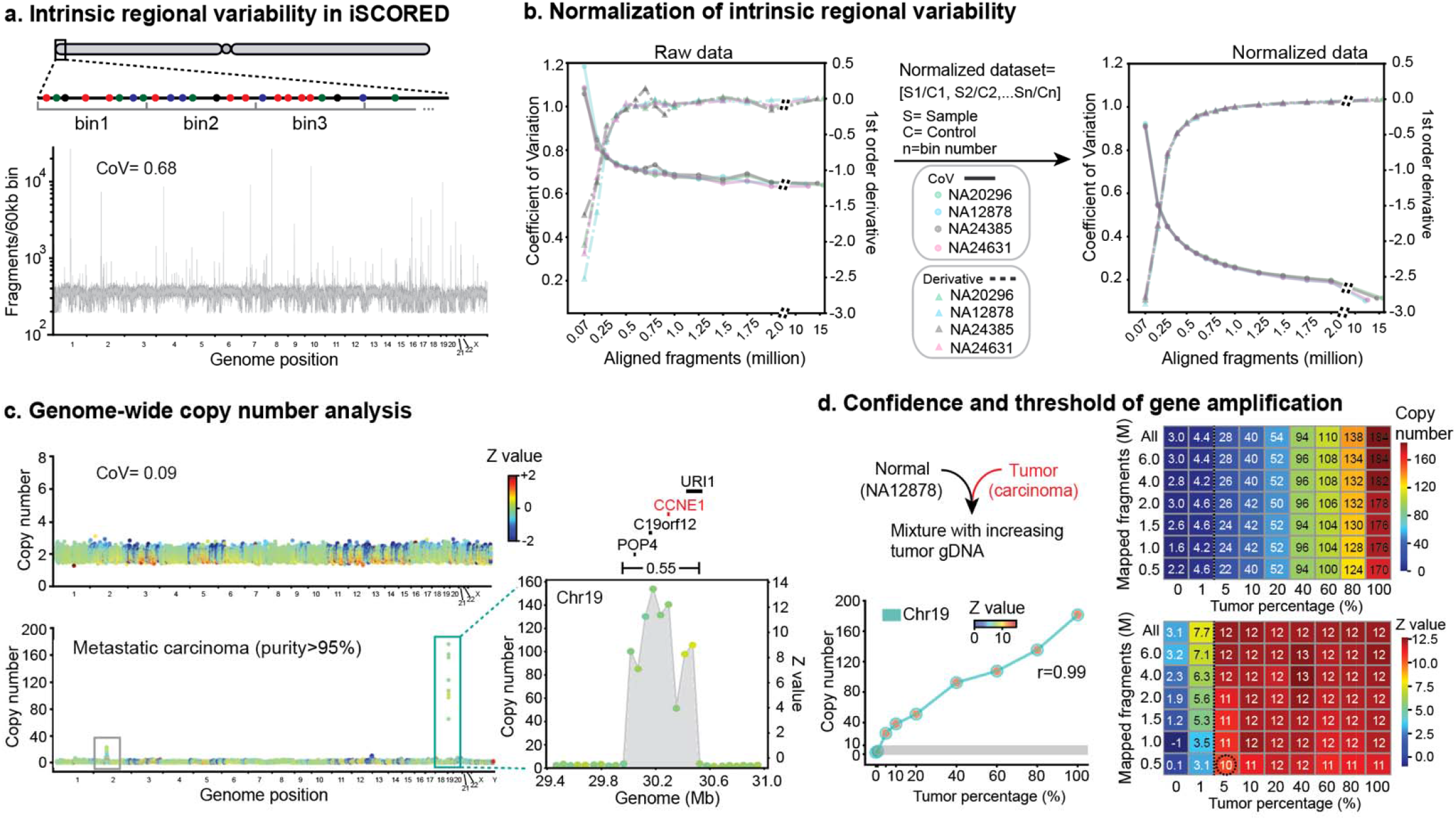
Normalization of variable mapped fragments in predefined bins for accurate copy number detection. **a**, The number of mapped fragments per bin fluctuates across the wild-type genome (intrinsic regional variability, IRV), yielding a relatively high coefficient of variation (CoV) of 0.68 and hampering detection of true outliers. **b**, Extensive sequencing does not address the fluctuation due to IRV (left panel). Normalizing the samples with the control wild-type dataset, the CoV dramatically drops and stabilizes at ∼1 million mapped fragments (right panel). **c**, The control genome data displayed CoV of 0.09 after normalization (upper panel). Application of this approach allows for detecting regions of amplification in both chromosome 2 and chromosome 19 (defined as copy number>10). **d**, Mixture of tumor with wild-type gDNAs shows that the amplified copies increase as the tumor percentage increases. Using Z values of 10 as cutoff, the genetic amplification *CCNE1* in chromosome 19 could be reliably detected at 5% tumor purity with 500,000 mapped fragments. Pearson Correlation of the data is shown.

We tested the top four overhang candidates that had the highest number of RE combinations while exhibiting the least possibility of methylation inhibition (Fig. 1c). To quantitatively measure the reconstruction efficiency (i.e., the number of uniquely mapped fragments per sequencing read), we compared combinations generating 4-nt overhangs with those generating 2-nt overhangs.

Surprisingly, we found that the efficiency of 4-nt overhang combinations is not superior to that of 2-nt overhang combinations (Fig. 1c, Figure S3). This is presumably due to the significantly higher number of generated fragments by REs with 2-nt overhangs. The most efficient combination was the TA overhang cocktail mix that consisted of MseI, BfaI, CviQI, and NdeI, resulting in a mean reconstruction efficiency of 4.6 and mapped fragment of 120 bp. To further optimize the iSCORED reaction, we tested various incubation periods and DNA ligases (Fig. 1d). Our experiments revealed that an incubation period of 30 minutes at 37 °C with intermittent agitation at 18 °C (900 rpm) yielded the highest mean reconstruction efficiency of 8.7 (Fig. 1d). This experimental condition was thus utilized for the remainder of the study.

### Genomewide aneuploidy detection in tumors

To detect large CNV (>10 Mb) and aneuploidy, the sequenced reads were first segmented into individual fragments by identifying short matches when mapping to the reference genome^21^. These uniquely mapped fragments were then filtered for quality (alignment scores ≥ 120, Methods for details) and assigned to predefined genomic bins (600 kb) for quantitative analysis. High numbers of mapped fragments per bin generated low variability between bins and this helped ensure high confidence in the resulting CNV plot. Finally, circular binary segmentation^24^ through DNACopy^25^ was employed to identify copy number alterations across genomic bins.

The performance of the iSCORED pipeline was compared to the conventional sequential SMURF approach^21^ (i.e. digestion, purification and ligation) and unprocessed native gDNA. By normalizing the datasets to the same amount of total DNA sequence, both the iSCORED and SMURF methods exhibited an over 16-fold increase in the number of fragments compared to native gDNA sequencing (Fig. 1e). The significant increase in fragment count resulted in low variability, which was critical for detecting copy number changes with high confidence. Specifically, the coefficient of variations (CoV) for the SMURF, iSCORED and native gDNA sequencing were 0.54, 0.57 and 3.3, respectively.

Detection of large CNV and aneuploidy by iSCORED showed 100% concordance with clinically validated chromosomal microarray data (Figure S9) and also demonstrated a much higher resolution than short-read based analysis^19^ within a comparable timeframe of 2 hours (Fig. 1f, Additional table 1). While the SMURF approach demonstrates a slightly better reconstruction efficiency (14.5), its sequential approach requires a substantial input DNA (2-3 μg)^21^ and an extended preparation time (90-120 minutes)^21,22^. This in contrast to the iSCORED method, which requires only 200-400 ng of input gDNA and a preparation time of 30 minutes. The prolonged process prohibits its intraoperative application in standard craniectomy surgeries (typically 3-4 hours; Additional table 2). The molecular information is crucial for accurate pathology diagnosis as many primary brain tumors are defined by molecular alterations. For instance, chromosomal 7 gains and 10 losses are characteristic of glioblastoma, while the presence of 1p/19q codeletion is required for oligodendroglioma diagnosis according to the 2021 WHO Classification^26^.

**Table 1:**
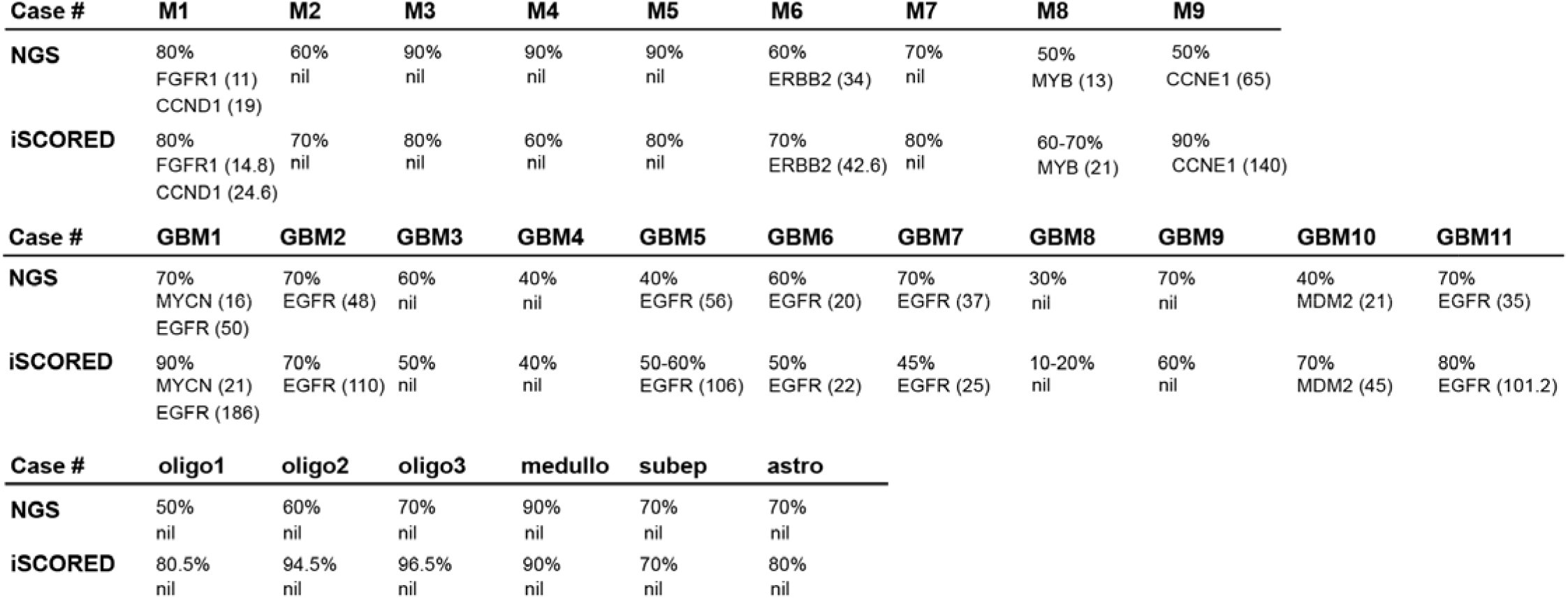
Comparison of CNV results between iSCORED and clinically validated assays. Percentage indicates the tumor purity of extracted DNA for corresponding assay. Numbers in the brackets are the detected copy number.

### Refinement of quantitative measures to detect copy number variations

While large 600kb bins are effective in detecting aneuploidy, most clinically-relevant gene amplifications occur within a range of hundreds of kilobases to a few megabases^27,28^. In such cases, using a large bin could result in averaging out the dose change, leading to decreased accuracy of small amplifications. Thus, we refined the bin size to 60 kb, which was similar to the highest genomic resolution of clinically validated chromosomal microarray analysis (CMA). When using the control human genome (NA12878) to quantify the total mapped fragments in refined 60-kb bins, the numbers mapped fragments fluctuated substantially across the predefined bins (Fig. 2a). Since the iSCORED is a restriction enzyme-based method, this finding was presumably due to variations in the density and distribution of restriction enzymes’ cutting sites across the human genome. We had termed the phenomenon intrinsic regional variability (IRV) (Fig. 2a, Figure S4). The background fluctuations might allow for tolerating outliers driven by true copy number changes, impacting the detection accuracy. In addition, this fluctuation behavior was inversely related to the amount of the data acquired (Fig. 2b). Hence, this finding was characterized in the context of the corresponding number of fragments in the genome.

For this purpose, we utilized the coefficient of variation (CoV)^29^ as a quantitative index, and performed time-lapsed analysis of the sequenced control samples. After comparing the sub-datasets with varying numbers of mapped fragments, we found that significant fluctuations reached a plateau around a CoV of 0.68 at one million mapped fragments, regardless of extensive sequencing (Fig. 2b, left panel). To address this issue, we performed bin-specific normalization by calculating a ratio of the mapped fragments in the sample of interest to those in the commonly used control reference genome (NA12878). This normalization significantly reduced the observed genomic fluctuations by approximately four-fold (Fig. 2b, right panel). The CoV of the normalized data was substantially reduced to 0.09 down from 0.68 in the corresponding non-normalized data.

This normalization process also allowed us to infer the required number of total mapped fragments to reliably identify regions of copy number change. When investigating the slope^30^ of CoV as a function of the acquired fragments, the inflection point was at a datapoint with mapped fragments of <500k, while the first order derivative function approached a value of zero (0) at about one million mapped fragments (Fig. 2b, right panel). Thus, we determined that acquiring approximately one million mapped fragments was sufficient to reliably detect genomic dosage changes. It is worth noting that pre-defined bins with inherently low counts can lead to high sampling variability, resulting false positive detections. Using normal well-characterized control gDNAs (NA20967, NA12878, NA24385 and NA24631 from Coriell Institute), we established that excluding bins within the lowest 0.2% genomic counts ensures reliable genomic dosage assessment (Figure S4). Bin-specific normalization and lowest 0.2% bin exclusion helped effectively detect true copy number variations by minimizing the effects of intrinsic regional variability.

### Gene amplification across various tumor purity levels

To demonstrate the effectiveness of the iSCORED procedure and its analysis pipeline, we first analyzed a metastatic adenosquamous carcinoma from esophagus (Case M9 in Table 1). A *CCNE1* gene amplification was detected in chromosome 19 (140 copies, inset in Fig. 2c), consistent with the molecular result from the clinically validated next generation sequencing. We further determined the minimum tumor percentage to reliably detect *CCNE1* amplification by assessing a range of mixtures comprising control gDNA (NA12878) and tumor gDNA. This revealed a positive correlation between increasing amplification and rising tumor percentage (Pearson *r* = 0.99). By utilizing a Z-score cutoff of 10 to establish detection confidence, we were able to detect *CCNE1* amplification in samples with as low as 5% tumor percentage using only 500k mapped fragments (Fig. 2d). Additionally, low copy number gain (22 copies) was also reliably detected with the same parameter, albeit at a higher tumor percentage and with more fragments (20% and 1.5 million fragments, respectively, Figure S5).

Overall, our results demonstrate the effectiveness of the iSCORED pipeline in detecting gene amplifications, even in samples with low tumor purity. The detection resolution outperforms the tumor percentage thresholds employed clinical next-generation sequencing platforms, which are generally set at 15-20%.

### Simulating intra-operative molecular analysis of the brain tumor cohort

We simulated the intraoperative molecular diagnosis by performing blind testing of a cohort of 26 intracranial neoplasms, including 17 primary CNS tumors and 9 metastatic tumors. Taking advantage of mechanical destruction of sectioning frozen tissue at 5 μm thickness using cryostat machine, high-quality gDNA is extracted within 15 minutes and processed through the iSCORED pipeline (see methods for details). The performance was timed and the findings were compared to the results from clinically validated next-generation sequencing (TruSight® Tumor 170 and whole exome sequencing) and chromosomal microarray analysis (Affymetrix OncoScan®)^23^.

Within one-hour of MinION sequencing, an average 344 ± 24 Mb of data were generated, corresponding to 1.38 ± 0.08 million mapped fragments (SEM, Figure S6). This is higher than the predetermined required data quantity for confident CNV detection (1×10^6^ mapped fragments, Fig. 2b). Furthermore, the output is approximately threefold the data volume of the recently published SMURF-based method (nCNV-seq) within the same 50-60 minutes sequencing window^22^. Across the 26 investigated samples, the diagnostic accuracy of iSCORED platform was 100% in detecting gene amplification of more than 10 copies (95% confidence interval^31^: 91%-100%, Pearson *r* = 0.81 by comparing to the NGS results; Table 1). One sample was detected to have *MYB* amplification (21 copies; case M8) by the iSCORED pipeline, a finding that was not originally uncovered by TST 170 panel but was later verified by a whole exome NGS study (13 copies).

The output genomic graph from the iSCORED pipeline provided precise information on amplified regions and the confidence of detected outliers (Fig. 2c and Figure S9). *EGFR* amplification is a molecular defining alterations in glioblastomas, typically occurring as extrachromosomal DNA ranging from 1-3 megabases (Mb) in size^27,32^. In our cohort of six *EGFR*-amplified glioblastomas, the average amplification regions spanned 1.66 ± 0.44 Mb (SEM) with an average copy number of 150.5 ± 47 (SEM). These samples also exhibited diverse regions and degrees of amplification, which is consistent with the known heterogeneity of glioblastoma^26,33^ (Fig. 3e).

**Fig. 3:**
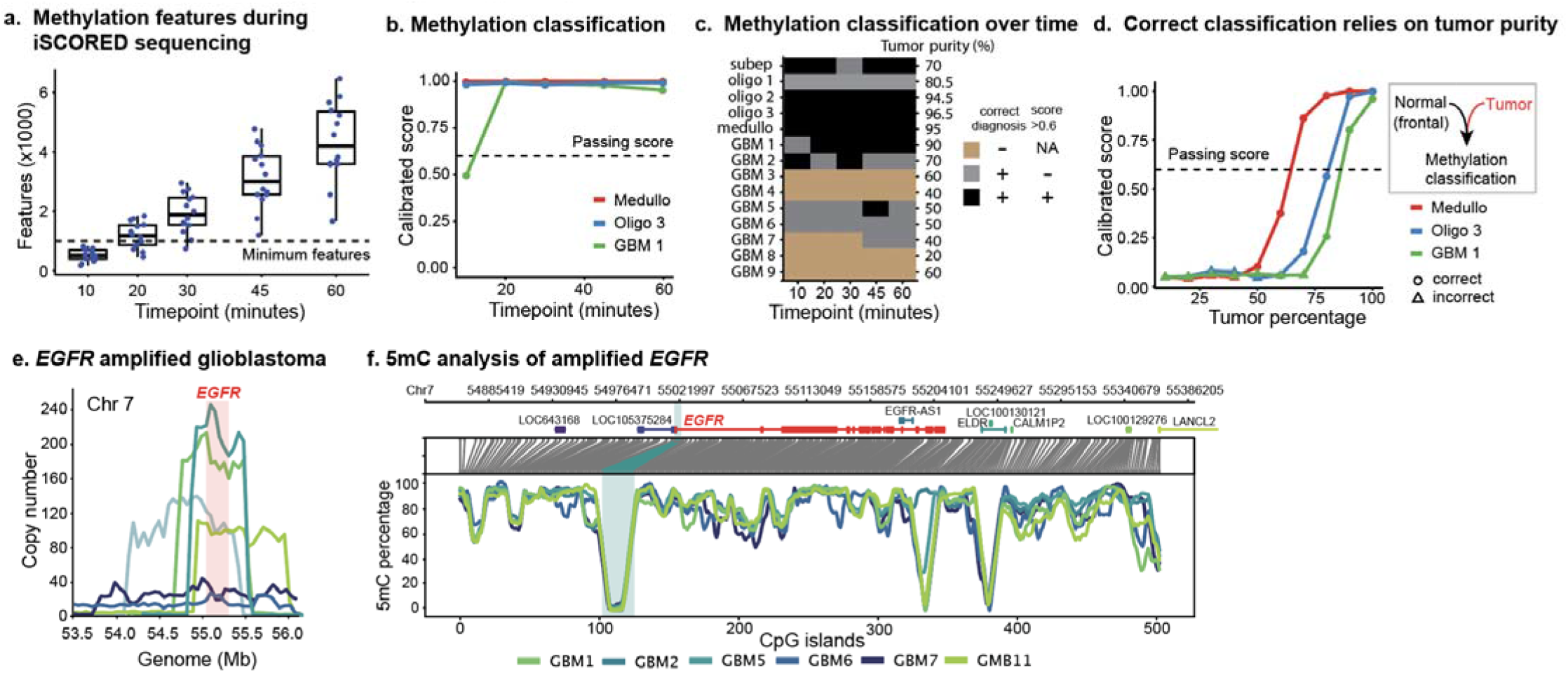
Simultaneous methylation analysis of primary CNV tumors. **a**, Minimal methylation classification features are acquired within 45 minutes of MinION sequencing. **b**, Calibrated methylation classification scores for glioblastoma, medulloblastoma and oligodendrogliomas with high tumor purity were calculated across multiple time points from the initiation of sequencing. **c**, Correct methylation classification of CNS tumors depends on high tumor purity. **d**, *In silico* mixture of glioblastoma, medulloblastoma and oligodendrogliomas with control brain tissue dataset at various ratios (total data quantity after one-hour of sequencing). **e**. Exact amplified regions covering EGFR oncogene in glioblastoma samples. **f.** Methylation characterization of amplified EGFR oncogene reveals promoter hypomethylation. Subep= subependymoma, oligo= oligodendroglioma, medullo= medulloblastoma, GBM= glioblastoma.

### Simultaneous methylation classification by iSCORED

Methylation classification of tumor types has emerged as an important diagnostic tool in clinical practice. Brain tumors in particular have benefited from the Heidelberg methylation classifier which classified 91 tumors across 2801 samples^34^. Given that ONT sequencing can identifying 5-methycytosine (5mC) from native DNA with no additional sample preparation, we extracted methylation information from our sequencing data and classified it with Rapid-CNS^2^, a machine learning-based classifier trained on the Heidelberg dataset^35^.

To evaluate the reliability of methylation classification over time, we processed MinION data at five timepoints (10, 20, 30, 45, and 60 minutes) and extracted the number of methylation features that overlapped with the 100k most variable features from the Heidelberg dataset (Fig. 3a). Within 45 minutes, all samples had identified more than 1000 CpG features, reaching the cut-off determined by the original authors^36^, and 10 out of 14 samples were correctly classified into tumor subclasses (Fig 3b, c and Figure S7). To investigate if the poor classification scores and misclassification were due to short fragments generated via iSCORED, we evaluated the data of three oligodendroglioma samples processed using iSCORED against the results from native DNA sequencing, as in the original publication^37^. The results revealed comparable classification scores (Figure S7c), indicating that fragmentation in iSCORED did not appear to affect the accuracy of methylation classification.

As the majority of glial neoplasms exhibit an infiltrative growth pattern, resected tumors are often mixed with normal brain parenchyma and inflammatory cells, which could significantly affect the methylation classification accuracy. This is supported by a positive correlation between histologically assessed tumor purity and classification score (Figure S7b). To further assess the impact of tumor purity on classification accuracy, we *in silico* admixed data from three tumors with the highest classification scores with control CNS tissue (frontal cortex, Fig. 3d). Our analysis revealed a rapid drop in classification scores as tumor purity decreased. Specifically, when the tumor purity was 60%, all classification scores fell below the commonly accepted classification threshold (0.6). Furthermore, when the tumor purity was below 40%, the samples were consistently assigned to incorrect classes. Notably, the medulloblastoma, which had the highest tumor purity, was least affected by control tissue mixing. Therefore, tumor purity is critical for successful tumor methylation classification.

### Promoter hypomethylation in the amplified oncogenes

The epigenetic landscapes of amplified oncogenes offer mechanistic insights into transcriptional regulations^27,38^. Despite the inherent low-pass nature of iSCORED platform, gene amplification ensures sufficient coverage for methylation profiling in the defined regions. Using glioblastoma as a proof of principle, within one-hour of MinION sequencing, we consistently detected hypomethylation across approximately 285 CpG sites within the promoters of the amplified *EGFR* (n=6, coverage depth of 7.6 ± 2.7 (SEM), Fig. 3e, f). Such promoter hypomethylation phenomenon is not exclusive to amplified *EGFR* of glioblastoma. Low 5mC percentages at CpG islands of oncogene promoters were detected in two other amplified oncogenes in glioblastomas (*MYCN* in GBM1 and *MDM2* in GBM6), as well as five oncogenes amplified in metastatic tumors (*FGFR1* and *CCND1* of breast cancer in M1, *ERBB2* of lung cancer in M6, *MYB* of esophageal cancer in M8 and *CCNE1* of esophageal cancer in M9; Figure S9).

### Timeframe for iSCORED pipeline

To facilitate intraoperative molecular diagnosis, an analysis pipeline was created that runs in conjunction with sequencing and finishes within minutes of sequencing completion. This pipeline includes a real-time basecalling process along with periodical filtering and alignment of samples for CNV, amplification, and methylation analysis. Once sufficient data is accumulated, the separate files are merged to finalize the analysis (Figure S8). The pipeline is designed to operate on standard computers, circumventing the need for complex and expensive infrastructure such as Cloud computing systems (see Methods for specific setup).

MinION sequencing typically requires 60 minutes to generate sufficient data. With the introduction of P2 Solo (ONT), four of our most recent specimens were analyzed using PromethION technology (R10.4.1). Remarkably, a mere 25 minutes of sequencing yielded 395 ± 55 Mb (SEM) of data on average, which corresponds to an average of 1.69 ± 0.37 million (SEM) mapped fragments (Figure S6a). Finally, the iSCORED platform and the real-time processing pipeline automatically generate a genome-wide copy number report and methylation classification within 5 minutes and 20 minutes of completing MinION and PromethION sequencing, respectively (140 and 120 minutes after receipt of specimen, Fig. 4). Thus, the iSCORED platform ensures an accurate, fast and inexpensive method for widespread clinical application.

**Fig. 4:**
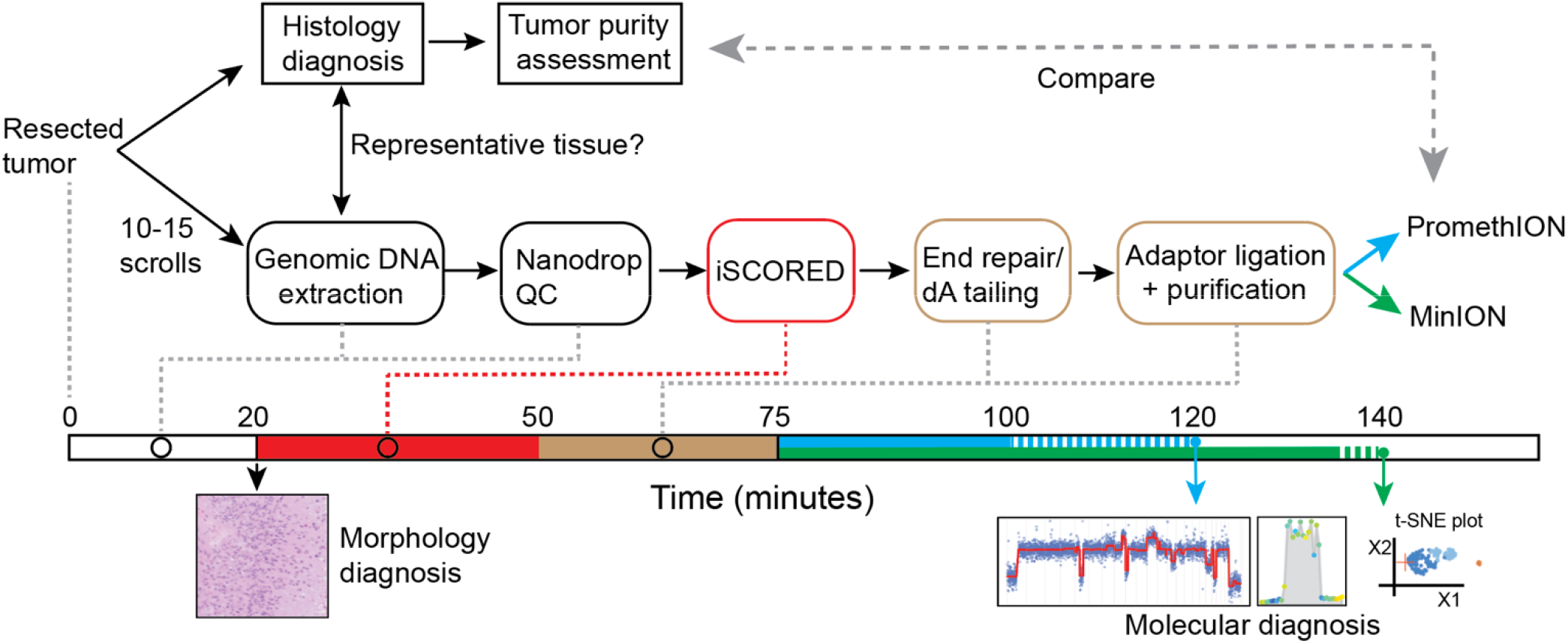
The iSCORED workflow for ultrafast molecular diagnosis. Shown is the incorporated iSCORED workflow applied during intraoperative morphology-based diagnosis. Additional 10-15 scrolls of tissue sections, at 5 μm thick, are prepared to extract gDNA for subsequent iSCORED library preparation. For achieving compatible sequencing results, two sequencing platforms are utilized: MinION, sequenced for 60 minutes with an additional 5 minutes of analysis, and PromethION, sequenced for 25 minutes with additional 20 minutes of data analysis (both with concurrent analysis during sequencing). The final output graphs comprise whole genome CNV, gene amplification regions and methylation classification with quantitative confidence scores (Z score for gene amplification and calibrated score for methylation classification).

### Cost-effectiveness

To reduce per-test costs, we evaluated the feasibility of re-using flowcells by using the ONT flow cell wash protocol (WSH004) after running each flow cell. Our results indicated that MinION flowcells can be re-used up to 7 times, before it dropped below the minimum requirement of 800 active pores. By employing the wash protocol, our results demonstrated 100% consistency with independent experiments using new flongle flowcells, indicating the absence of detectable carryover between experiments (Figure S9). The ability to reuse flow cells resulted in a reduced sequencing cost of $125 per sample (Additional table 3).

## Discussion and conclusion

In this study, we present iSCORED, a novel library preparation method that can rapidly and affordably generate high resolution CNV profiles, detect gene amplifications, and classify tumors by their methylation status. While Nanopore has well known capabilities in long-read sequencing, it is not inherently optimized for shorter reads due to the potential delay on molecules reloading and unloading to sequencing pores. We overcome this limitation by employing iSCORED method to concatenate small digested genomic fragments in one reaction, significantly improving sequencing efficiency and achieving molecular analysis within craniotomy surgical windows (3-4 hours). In contrast to other sequencing platforms^17^ and hybridization-based arrays^23,39^, the low cost per sample, a mere $125 USD, and the ease of setting up the infrastructure with a budget of $6,000-8,000 USD for MinION and $14,000-16,000 USD for PromethION make it an economical option for clinical applications (Additional table 3). Notably, the unmatched turnaround time of 120-140 minutes further positions our method as a robust and invaluable tool for widespread implementation in clinical settings (Additional table 1).

The 2021 WHO classification of the Central Nervous System Tumors has incorporated a substantial amount of molecular diagnostic alterations for accurate classification of CNS tumors^26^. Genome-wide copy number analysis and methylation profiling are imperative for proper tumor grading and integrated diagnoses. The iSCORED platform demonstrates a high accuracy in detecting gene amplifications, with the thresholds set at 5-20% of tumor purities for high and low copy number amplifications, respectively. Concurrent 5-mC information could be obtained for tumor methylation profiling. However, it is important to note that in the methylation classification of primary CNS tumors, the sensitivity related to tumor purity becomes apparent. Using a calibrated score of 0.6 as the cutoff, accurate classification typically requires a tumor percentage of more than 60%. The phenomenon is not unique to iSCORED platform but also present in other DNA methylation arrays^34^. It is thus crucial to assess the tissue quality and estimate the tumor percentage during the morphology-based intraoperative diagnosis (Fig. 4). Nevertheless, the iSCORED platform offers an ultrafast and comprehensive approach to characterizing CNV and conducting tumor methylome profiling.

We have shown that CpG methylation can be measured within the amplified regions. Previous studies involving cell lines indicated that oncogene promoters, when presented as extrachromosomal DNA, exhibited lower methylation compared to the chromosomal DNA of the same gene loci^27^. In our cohort of 26 snap-frozen patient tumors, we identified eight distinct oncogene amplifications, all of which exhibited hypomethylated promoters. The pattern suggests epigenetic modifications could serve as a fundamental mechanism for active transcription in the amplified oncogenes. As a result, a combined approach integrating both genetic (amplification) and epigenetic (promoter methylation) data could be a better parameter for predicting protein expression, tumor behavior and clinical outcomes.

Somatic copy number alterations represent a major type of genetic mutations in cancer initiation, progression and treatment resistance^1–3^. Among the various cancer types, ovarian carcinoma and sarcoma bear the highest burden of CNVs, accounting for approximately 80% of cases. Following closely are uterine carcinosarcoma and esophageal carcinomas, with approximately 75% exhibiting CNVs^40^. These findings underscore the importance of CNV analysis and the applicability of the iSCORED platform in diverse cancer types. Accurate identification and comprehensive understanding of the prevalence and implications of CNVs in these cancers are pivotal for advancing diagnostic accuracy and prognostic evaluation. Moreover, ultrafast detection of oncogene amplification would augment molecular targeted therapeutics. For instance, Trastuzumab (Herceptin) is a monoclonal antibody that binds to the HER2 receptor and is employed to treat *ERBB2*-amplified breast and gastric cancers^41,41–45^. Several EGFR inhibitors are used to treat *EGFR*-amplified cancers, such as gefitinib and erlotinib for non-small cell lung cancer^46,47^, and cetuximab for colorectal cancer^48^. This rapid molecular characterization can expedite the prompt administration of molecular targeted drugs during or early after surgery, resulting in superior therapeutic effects. Therefore, integrating iSCORED-based ultrafast molecular diagnosis into the therapeutic strategies could unlock a new frontier in personalized medicine.

## Methods

### DNA extraction from OCT embedded frozen tissue

The study was approved by the Institutional Review of Board (IRB) of Dartmouth-Hitchcock Medical Center (DHMC, STUDY02001960). The banked samples were retrieved from the institutional biorepository at the DHMC. Genomic DNA was extracted using the DNeasy Blood & Tissue Kit (Qiagen #69504) with minor modifications for ultrafast extraction. Briefly, 5-10 scrolls of tissue (>5mm x 5mm) were sectioned at 5 μm thickness onto blank slides in a cryostat machine. A premixed solution containing 180 μl of tissue lysis buffer (buffer ATL) and 2 μl of RNase A (Thermo Scientific #01236994) was added onto the slide and the tissue was scrapped off, then transferred to a 1.5 ml Eppendorf tube. The tube was incubated at 37 °C for 2 minutes, after which 20 μl of protease K was added to the reaction, followed by an additional incubation at 56 °C for 8 minutes. Buffer AL (200 μl) and pure ethanol (200 μl) were then subsequently added to the reactions. The final reaction was mixed thoroughly by vortexing and added to the spin column inserted to the vacuum to facilitate the extraction procedures. Sequential buffer AW1 (500 μl) and AW2 (500 μl) were added to wash the column, which was centrifuged at 20,000 x g for 30 seconds for final cleanup. The DNA was eluted with 50-75 μl of AE buffer and its quantity and quality were checked using Nanodrop and Qubit instruments (ThermoFisher).

### iSCORED Reaction

Approximately 200-400 ng of input gDNA was used for the iSCORED reaction, followed by bead purification for Nanopore sequencing. The reaction mixture comprised 15 μl in total, including quick ligase buffer, quick ligase (NEB E6056) and NTAN cocktail mix (NEB) and extracted genomic DNA. The reaction was incubated at 37 °C for 30 minutes with intermittent cooling/agitation to enhance ligation. End repair/dA tailing buffer and enzyme mix (NEB E7546) were added to the mixture, which was then incubated at 20 °C for 5 minutes and 65 °C for 5 minutes. For final ligation to the Nanopore motor proteins, a freshly made premix of ligation buffer (LNB, 4 μl), adaptor protein (AMF, 1.5 μl, ONT LSK110 for R9.4.1 flowcells; LA 1.5 μl ONT LSK114 for R10.4.1 flowcells) and quick ligase (1.5 μl) were added to the reaction solution and incubated for 10 minutes at room temperature (20-22 °C).

Finally, AMPure XP beads (Beckman Coulter A63881) were added for standard magnetic bead purification. The beads were washed twice with long fragment buffer (LFB, 80 μl) before eluting into 12 μl elution buffer. Approximately 30-50 ng of ligated DNA were loaded for flongle flowcells (R9.4.1), 100 ng were used for MinION flowcells (R9.4.1) and 50-75 ng for PromethION flowcells (R10.4.1).

### Flowcell reuse

The MinION (R9.4.1) and PromethION (R10.4.1) flowcells were washed for sequential runs by using the flowcell wash kit (WSH004-XL). Briefly, 400 μl of flowcell wash mix (398 μl of wash diluent and 2 μl of wash mix) were loaded to the priming port to allow a DNase I reaction for 60 minutes at room temperature. The reaction solution was removed from the waste port and storage buffer (500 μl) were loaded into the priming port before storing at 4 °C for next use. A minimum of 800 active pores by flowcell check were required for a successful run. Following the protocol, a typical MinION flowcell (R9.4.1) could be re-used for 5-7 times.

### Normalization of intrinsic regional variability

Control human genomic DNA (gDNA; NA12878 from Coriell Institute) was first processed with iSCORED and sequenced extensively (16,564,873 mapped fragments) to establish a reference dataset for normalization to equivalent bins. The bins with counts below 0.2^th^ percentile were removed to eliminate false positive bins as tested in four control datasets (NA20967, NA12878, NA24385 and NA24631 from Coriell Institute; Figure S4). An array of the proportion was segmented at the chromosomal level to optimize the resolution of the normalization and increase the sensitivity at which outliers are detected. A normalized vector of ratios (r1), identical in size to the reference array, was created. The distribution of this vector, at the chromosomal level, was employed to detect outliers using three components: i) a threshold of 5 to filter the elements in r1 that were not greater than a specific threshold =5, ii) Z scores to determine the statistical significance of the deviation from the distribution, and iii) the presence of surrounding outlier bins (a minimum of two consecutive bins must be present for a set of datapoints to be considered as outliers).

### Coefficient of variation calculation across genome

Similar to what was described above, a normalized vector was created using NA12878 as a reference. For this approach, we used three other commonly used control gDNAs (NA20296, NA24385 and NA24631 from Coriell Institute). Each one of these datasets was segmented into independent data subsets (with no overlapping fragments) that vary in the number of mapped fragments. The number of fragments in these datasets were 70k, 200k, 300k, 400k, 500k, 600k, 700k, 800k, 900k, 1M, 1.25M, 1.75M, 2M. For each datasets, the coefficient of variation (CoV) across the genome was calculated to assess the variability. In addition, the behavior of the CoV as the number of fragments change was assessed with the first order derivative of the CoV function.

### Basecalling and read filtering

Fast5 files were converted to pod5 with ONT’s pod5-file-format (https://github.com/nanoporetech/pod5-file-format) and basecalled with ONT’s Dorado v0.2.4 (https://github.com/nanoporetech/dorado) using dna_r9.4.1_e8_fast@v3.4 for r9.4.1 or dna_r10.4.1_e8.2_400bps_sup@v3.5.2 for r10.4.1 with the following setting:--modified-bases 5mCG--emit-moves. The resulting unmapped SAM (uSAM) files were converted to fastq using samtools fastq-TMM, ML to carry the methylation information forward into the fastq header. The resulting FastQs were first processed with Porechop (v0.2.1, https://github.com/rrwick/Porechop) to trim adapter sequences and split reads with internal adapters. Filtered with NanoFilt (2.8.0) to remove rare reads greater than 15kb which represented native genomic reads that did not contribute to our analysis^49^.

### Read processing into aligned fragments

The following was adapted from Prabakar et al.^50^. Briefly, filtered reads were aligned to GRCh37/hg19 using bwa-mem^51^ (v0.7.17) with the following settings: -x ont2d -k 12 -W 12 -A 4 -B 10 -O 6 -E 3 -T 120. These settings allowed the segmentation of the concatenated reads into individual fragments that were aligned to their respective genomic regions. To ensure accurate quantitative CNV analysis, the duplex reads were identified and excluded from the original uSAM with ONT-Duplex Tools (https://github.com/nanoporetech/duplex-tools). A single member of each pair was then removed from the SAM file using a list of readIDs and Picard (https://github.com/broadinstitute/picard). The genome was then subdivided into either 5,000 or 50,000 genomic bins for CNV and amplification analysis, respectively, and mapped fragments per bin were calculated.

### CNV analysis

CNV analysis was performed using the Smurf-seq analysis pipeline^50^. Counts of uniquely mapped fragments to the 5,000 bins in the human genome were normalized for biases in GC content, finally an implementation of DNAcopy^52^ (v1.74.1) using circular binary segmentation identified breakpoints in bin counts.

### Output table and graph

The output table contained a list of at least two consecutive statistically significant outliers (i.e., bins) to minimize the potential of identifying isolated/noisy outliers due to individual genome variation. The table displayed the corresponding position, ratio, Z score at the chromosomal level of each sample, along with commonly amplified gene(s) found in these bins of interest. All annotated genes from the hg19 reference genome were included in the table with 75 commonly amplified genes highlighted in red. The graph was automatically generated if there are significant bins in the sample of interest.

### Methylation calling and tumor classification

The following were adapted from Rapid-CNS^2^ pipeline^37^. The filtered reads were aligned to GRCh38/hg38 using bwa-mem^51^ (v0.7.17) with the following settings: -x ont2d -k 12 -W 12 -A 4 -B 10 -O 6 -E 3 -T 120 -C -Y to map the individual fragments within each read. The -C command allowed the SAM tags for methylation to be moved from the fastq header back into the SAM file. The -Y command turned off soft-clipping which would otherwise de-couple the methylation tag information. The per site methylation is extracted using mbtools (https://github.com/jts/mbtools). A custom python script converted the bedfile to make it compatible with Rapid-CNS^2^ which processes the methylation information using a random forest classifier trained on Illumina BeadChip 450 K methylation array from the Heidelberg reference cohort of brain tumor methylation profiles^53^.

To determine the minimum time required for methylation classification, we simulated the collection of methylation data over time using samples that had been sequenced for >60 minutes. The data were subdivided into several bins (10, 20, 30, 45, 60 minutes). Sequencing start time was recovered from the uSAM read header and the aligned SAM was filtered accordingly. The data was then processed using our standard analysis pipeline as described above to extract the number of detected methylation features, the methylation classification, and the calibrated scores at each time point.

To study the role of tumor percentage in methylation classification, gDNA from control human frontal lobe was processed with iSCORED. The resulting reads were *in silico* admixed with datasets from a medulloblastoma, an oligodendroglioma, or a glioblastoma, all of which had >90% tumor percentage and calibrated scores of ∼0.99 in methylation classification. Reads equivalent to an hour of sequencing on the MinION at different ratios of tumor to control (tumor percentages of: 0-100 in intervals of 10) were used for methylation classification using the Rapid-CNS^2^ pipeline.

### Computer setting

Our computer system included an Intel® Core™ i9-12900K Processor, 24 cores, 64 GB of RAM, 2Tb of storage and an RTX 3090Ti.

### Chromosomal Microarray (Affymetrix OncoScan®)

DNA from FFPE samples was isolated using the QIAGEN QIAamp FFPE Tissue Kit (Qiagen, Valencia, CA). DNA quantity was measured with the Qubit Fluorometer 3.0 and Qubit dsDNAHigh-Sensitivity assay kit (Thermo Fisher Scientific company, Waltham, MA). Samples were then subjected to CMA following the protocol of the OncoScan FFPE Assay Kit (Affymetrix, Santa Clara, CA) described previously^23^.

### Next Generation Sequencing

Tissue samples used in this study had been previously sequenced using either the Illumina TruSight® Tumor 170 assay or the newer DHCancerSeq whole exome sequencing assay. Both tests had been clinically validated in the Center for Clinical Genomics and Advanced Technology at the Dartmouth Hitchcock Medical Center Department of Pathology and Laboratory Medicine for routine clinical use. Each assay was validated to perform NGS using DNA and RNA isolated from formalin-fixed, paraffin-embedded (FFPE) tissue samples for somatic analysis with an integrated bioinformatics pipeline for sequencing analysis, variant calling, and interpretation. The same set of 170 genes were analyzed regardless of which assay was used on the samples for this study. For the DHCancerSeq, the Agilent SureSelect Human All Exon V8 was used to provide a comprehensive and most up-to-date coverage of protein coding regions from RefSeq, CCDS, and GENCODE. The SureSelect Human All Exon V8 spans a 35.1 Mb target region of the human genome with an end-to-end design size of 41.6 Mb. The V8 exome workflow is automated with the Magnis NGS Prep System. The Illumina TST170 workflow was automated using the Beckman Coulter Biomek NX^P^ robotic workstations. Each assay was designed to examine single nucleotide variants, small deletions, small insertions, amplifications, fusions, and splice site variants to obtain a comprehensive somatic molecular profile for diagnosis, prognosis, and prediction of therapeutic response. TST170 libraries were sequenced on the Illumina NexSeq500 and DHCancerSeq libraries sequenced on the Illumina NovaSeq 6000 System. RNA library preparations and target enrichment were performed using the Illumina TST-170 sequencing assay.

Routine clinical NGS was performed by either the Illumina TST170 assay or the DHCancerSeq which utilizes the Agilent SureSelect Human Exome V8. The same 170 gene targets were evaluated in each assay.

TST 170 amplification targets:

*AKT2, ALK, AR, ATM, BRAF, BRCA1, BRCA2, CCND1, CCND3, CCNE1, CDK4, CDK6, CHEK1, CHEK2, EGFR, ERBB2,ERBB3, ERCC1, ERCC2, ESR1, FGF1, FGF10, FGF14, FGF19, FGF2, FGF23, FGF3, FGF4, FGF5, FGF6, FGF7, FGF8, FGF9, FGFR1, FGFR2, FGFR3, FGFR4, JAK2, KIT, KRAS, LAMP1, MDM2, MDM4, MET, MYC, MYCL1, MYCN, NRAS, NRG1, PDGFRA, PDGFRB, PIK3CA, PIK3CB, PTEN, RAF1, RET, RICTOR, RPS6KB1, TFRC*.

### Methylation profiling of amplified oncogenes

Methylation information from the bam files were plotted using MethylArtst^54^ using the ‘region’ function with a setting of 1000 windows and a smoothing window of 4. Gene and promoter locations for important oncogenes were extracted from Ensembl for GRCh38. A region spanning 0.8x of the length of the gene upstream and downstream was plotted, highlighting the promoter region.

## Declarations

### Availability of data and materials

The iSCORED analysis pipeline for processing raw sequencing data will be available upon publication of the paper.

## Author contributions

Conceived iSCORED: CCL. Designed the experiments: FEE, AOO, and CCL. Performed the experiments: FEE, AOO, and CCL. Analyzed the data: FEE, AOO, and CCL. Histology analysis: GJZ. Contributed Oncoscan and NGS results: EGH and GJT. Wrote the paper: FEE, AOO, and CCL. All authors read, edited and contributed to the final version of the manuscript.

## Acknowledgements

We thank Sandeep Wontakal and Eric Loo for comments on the manuscript and discussions. We thank Florian Schroeck for helping with calculating the one-tailed confidence interval for the detection accuracy of the method. We are grateful to Rachael Barney and Torrey Gallagher for retrieval of biospecimens and experimental support. The authors acknowledge the support of the Laboratory for Clinical Genomics and Advanced Technology in the Department of Pathology and Laboratory Medicine of the Dartmouth Hitchcock Health System.

## References

1. Beroukhim, R. et al. The landscape of somatic copy-number alteration across human cancers. Nature 463, 899–905 (2010).

2. Zack, T. I. et al. Pan-cancer patterns of somatic copy number alteration. Nat. Genet. 45, 1134– 1140 (2013).

3. Stratton, M. R., Campbell, P. J. & Futreal, P. A. The cancer genome. Nature 458, 719–724 (2009).

4. Levchenko, A., Kanapin, A., Samsonova, A. & Gainetdinov, R. R. Human Accelerated Regions and Other Human-Specific Sequence Variations in the Context of Evolution and Their Relevance for Brain Development. Genome Biol. Evol. 10, 166–188 (2018).

5. Dumas, L. et al. Gene copy number variation spanning 60 million years of human and primate evolution. Genome Res. 17, 1266–1277 (2007).

6. Marques-Bonet, T. et al. A burst of segmental duplications in the genome of the African great ape ancestor. Nature 457, 877–881 (2009).

7. Heinzen, E. L. et al. Genome-wide scan of copy number variation in late-onset Alzheimer’s disease. J. Alzheimers Dis. JAD 19, 69–77 (2010).

8. Lee, W.-P. et al. Copy Number Variation Identification on 3,800 Alzheimer’s Disease Whole Genome Sequencing Data from the Alzheimer’s Disease Sequencing Project. Front. Genet. 12, 752390 (2021).

9. Hastings, P. J., Lupski, J. R., Rosenberg, S. M. & Ira, G. Mechanisms of change in gene copy number. Nat. Rev. Genet. 10, 551–564 (2009).

10. Pinto, D. et al. Convergence of genes and cellular pathways dysregulated in autism spectrum disorders. Am. J. Hum. Genet. 94, 677–694 (2014).

11. Pinto, D. et al. Functional impact of global rare copy number variation in autism spectrum disorders. Nature 466, 368–372 (2010).

12. Malhotra, D. & Sebat, J. CNVs: harbingers of a rare variant revolution in psychiatric genetics. Cell 148, 1223–1241 (2012).

13. Kallioniemi, A. et al. Comparative genomic hybridization for molecular cytogenetic analysis of solid tumors. Science 258, 818–821 (1992).

14. Wang, D. G. et al. Large-scale identification, mapping, and genotyping of single-nucleotide polymorphisms in the human genome. Science 280, 1077–1082 (1998).

15. Pinkel, D. et al. High resolution analysis of DNA copy number variation using comparative genomic hybridization to microarrays. Nat. Genet. 20, 207–211 (1998).

16. Ciriello, G. et al. Emerging landscape of oncogenic signatures across human cancers. Nat. Genet. 45, 1127–1133 (2013).

17. Malone, E. R., Oliva, M., Sabatini, P. J. B., Stockley, T. L. & Siu, L. L. Molecular profiling for precision cancer therapies. Genome Med. 12, 8 (2020).

18. Gorzynski, J. E. et al. Ultrarapid Nanopore Genome Sequencing in a Critical Care Setting. N. Engl. J. Med. 386, 700–702 (2022).

19. Wei, S. et al. Rapid Nanopore Sequencing-Based Screen for Aneuploidy in Reproductive Care. N. Engl. J. Med. 387, 658–660 (2022).

20. Wang, Z. et al. SMASH, a fragmentation and sequencing method for genomic copy number analysis. Genome Res. 26, 844–851 (2016).

21. Prabakar, R. K., Xu, L., Hicks, J. & Smith, A. D. SMURF-seq: efficient copy number profiling on long-read sequencers. Genome Biol. 20, 134 (2019).

22. Wongsurawat, T. et al. Exploiting nanopore sequencing for characterization and grading of IDH-mutant gliomas. Brain Pathol. Zurich Switz. e13203 (2023) doi:10.1111/bpa.13203.

23. Jung, H.-S., Lefferts, J. & Tsongalis, G. Utilization of the oncoscan microarray assay in cancer diagnostics. Appl. Cancer Res. 37, (2017).

24. Olshen, A. B., Venkatraman, E. S., Lucito, R. & Wigler, M. Circular binary segmentation for the analysis of array-based DNA copy number data. Biostat. Oxf. Engl. 5, 557–572 (2004).

25. Seshan, V. E. & Olshen, A. DNAcopy: DNA copy number data analysis.

26. Louis, D. N., et al. The 2021 WHO Classification of Tumors of the Central Nervous System: a summary. Neuro-Oncol. 23, 1231–1251 (2021).

27. Hung, K. L. et al. Targeted profiling of human extrachromosomal DNA by CRISPR-CATCH. Nat. Genet. 54, 1746–1754 (2022).

28. Helmsauer, K. et al. Enhancer hijacking determines extrachromosomal circular MYCN amplicon architecture in neuroblastoma. Nat. Commun. 11, 5823 (2020).

29. Virtanen, P. et al. SciPy 1.0: fundamental algorithms for scientific computing in Python. Nat. Methods 17, 261–272 (2020).

30. Quarteroni, A., Sacco, R. & Saleri, F. Numerical Mathematics. (Springer New York Springer e-books Imprint: Springer, 2007).

31. Newcombe, R. G. Interval estimation for the difference between independent proportions: comparison of eleven methods. Stat. Med. 17, 873–890 (1998).

32. Verhaak, R. G. W., Bafna, V. & Mischel, P. S. Extrachromosomal oncogene amplification in tumour pathogenesis and evolution. Nat. Rev. Cancer 19, 283–288 (2019).

33. Parker, N. R., Khong, P., Parkinson, J. F., Howell, V. M. & Wheeler, H. R. Molecular heterogeneity in glioblastoma: potential clinical implications. Front. Oncol. 5, 55 (2015).

34. Capper, D. et al. DNA methylation-based classification of central nervous system tumours. Nature 555, 469–474 (2018).

35. Patel, A. et al. Rapid-CNS2: rapid comprehensive adaptive nanopore-sequencing of CNS tumors, a proof-of-concept study. Acta Neuropathol. (Berl.) 143, 609–612 (2022).

36. Euskirchen, P. et al. Same-day genomic and epigenomic diagnosis of brain tumors using real-time nanopore sequencing. Acta Neuropathol. (Berl.) 134, 691–703 (2017).

37. Patel, A. et al. Rapid-CNS2: rapid comprehensive adaptive nanopore-sequencing of CNS tumors, a proof-of-concept study. Acta Neuropathol. (Berl.) 143, 609–612 (2022).

38. Siegfried, Z. et al. DNA methylation represses transcription in vivo. Nat. Genet. 22, 203–206 (1999).

39. Noguera-Castells, A., García-Prieto, C. A., Álvarez-Errico, D. & Esteller, M. Validation of the new EPIC DNA methylation microarray (900K EPIC v2) for high-throughput profiling of the human DNA methylome. Epigenetics 18, 2185742 (2023).

40. Harbers, L. et al. Somatic Copy Number Alterations in Human Cancers: An Analysis of Publicly Available Data From The Cancer Genome Atlas. Front. Oncol. 11, 700568 (2021).

41. Modi, S. et al. Trastuzumab Deruxtecan in Previously Treated HER2-Positive Breast Cancer. N. Engl. J. Med. 382, 610–621 (2020).

42. Piccart-Gebhart, M. J. et al. Trastuzumab after adjuvant chemotherapy in HER2-positive breast cancer. N. Engl. J. Med. 353, 1659–1672 (2005).

43. Romond, E. H. et al. Trastuzumab plus adjuvant chemotherapy for operable HER2-positive breast cancer. N. Engl. J. Med. 353, 1673–1684 (2005).

44. Slamon, D. J. et al. Use of chemotherapy plus a monoclonal antibody against HER2 for metastatic breast cancer that overexpresses HER2. N. Engl. J. Med. 344, 783–792 (2001).

45. Shitara, K. et al. Trastuzumab Deruxtecan in Previously Treated HER2-Positive Gastric Cancer. N. Engl. J. Med. 382, 2419–2430 (2020).

46. Cataldo, V. D., Gibbons, D. L., Pérez-Soler, R. & Quintás-Cardama, A. Treatment of non-small-cell lung cancer with erlotinib or gefitinib. N. Engl. J. Med. 364, 947–955 (2011).

47. Yuan, M., Huang, L.-L., Chen, J.-H., Wu, J. & Xu, Q. The emerging treatment landscape of targeted therapy in non-small-cell lung cancer. Signal Transduct. Target. Ther. 4, 61 (2019).

48. Jonker, D. J. et al. Cetuximab for the treatment of colorectal cancer. N. Engl. J. Med. 357, 2040–2048 (2007).

49. De Coster, W., D’Hert, S., Schultz, D. T., Cruts, M. & Van Broeckhoven, C. NanoPack: visualizing and processing long-read sequencing data. Bioinforma. Oxf. Engl. 34, 2666–2669 (2018).

50. Prabakar, R. K., Xu, L., Hicks, J. & Smith, A. D. SMURF-seq: efficient copy number profiling on long-read sequencers. Genome Biol. 20, 134 (2019).

51. Li, H. & Durbin, R. Fast and accurate short read alignment with Burrows-Wheeler transform. Bioinforma. Oxf. Engl. 25, 1754–1760 (2009).

52. Venkatraman E. Seshan, A. O. DNAcopy. (2017) doi:10.18129/B9.BIOC.DNACOPY.

53. Kuschel, L. P. et al. Robust methylation-based classification of brain tumours using nanopore sequencing. Neuropathol. Appl. Neurobiol. (2022) doi:10.1111/nan.12856.

54. Cheetham, S. W., Kindlova, M. & Ewing, A. D. Methylartist: tools for visualizing modified bases from nanopore sequence data. Bioinforma. Oxf. Engl. 38, 3109–3112 (2022).

