## Supplementary data for "iSCORED: nanopore-based random genomic sampling for intraoperative molecular diagnosis"

Additional table 1: Comparison of CNV detection methods

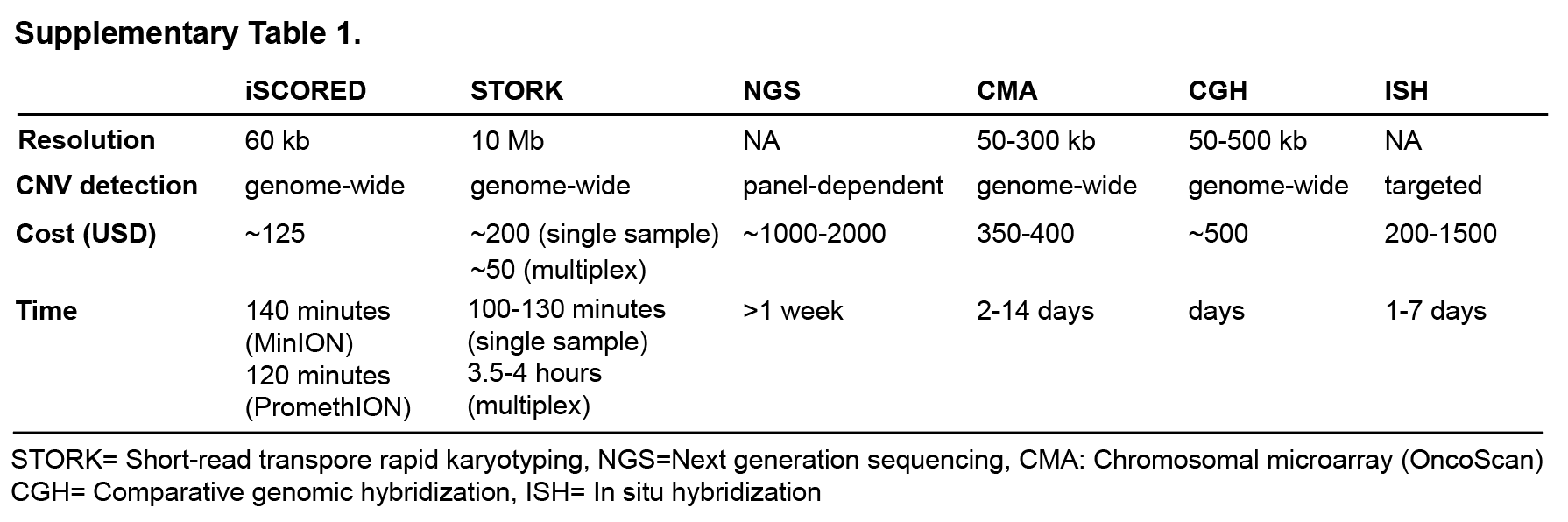

Additional table 2: Comparison of iSCORED and SMURF methods

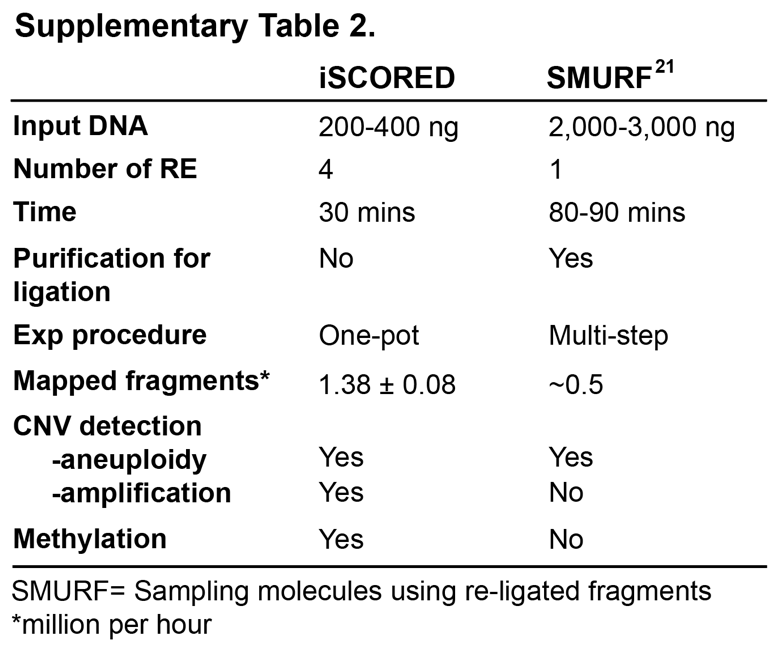

Additional table 3: Reagent and setup fee for iSCORED platform

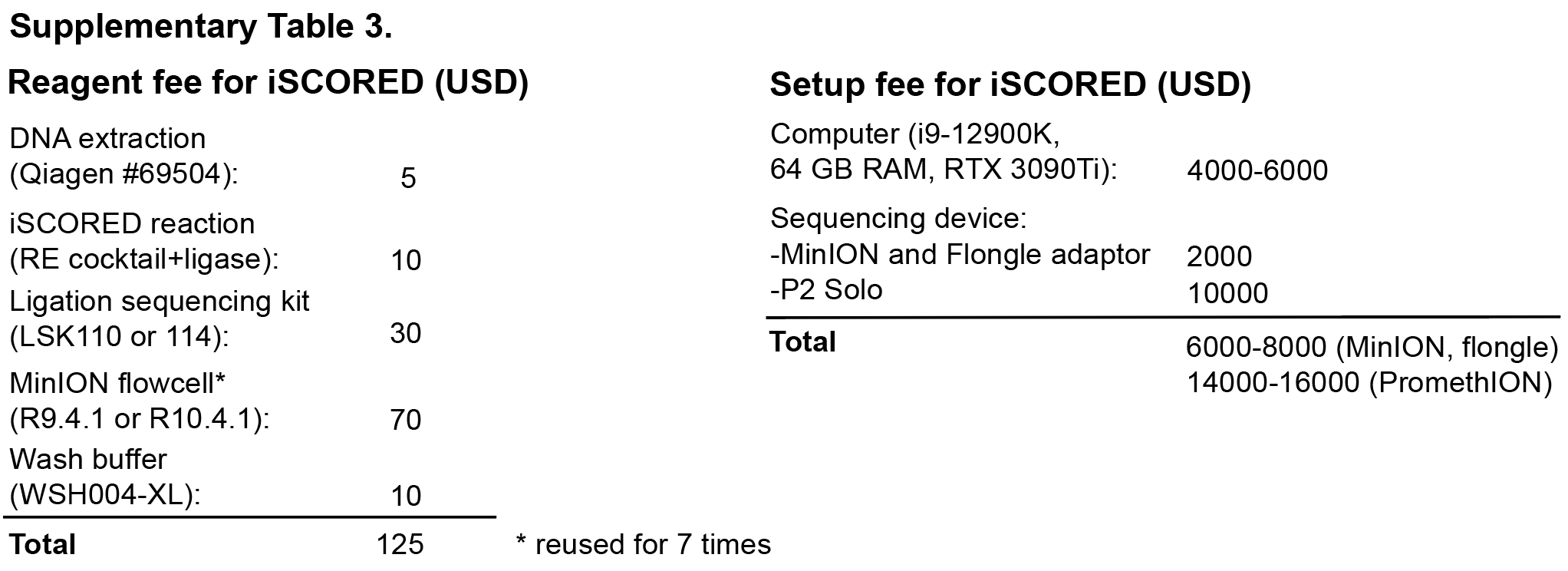

Figure S1

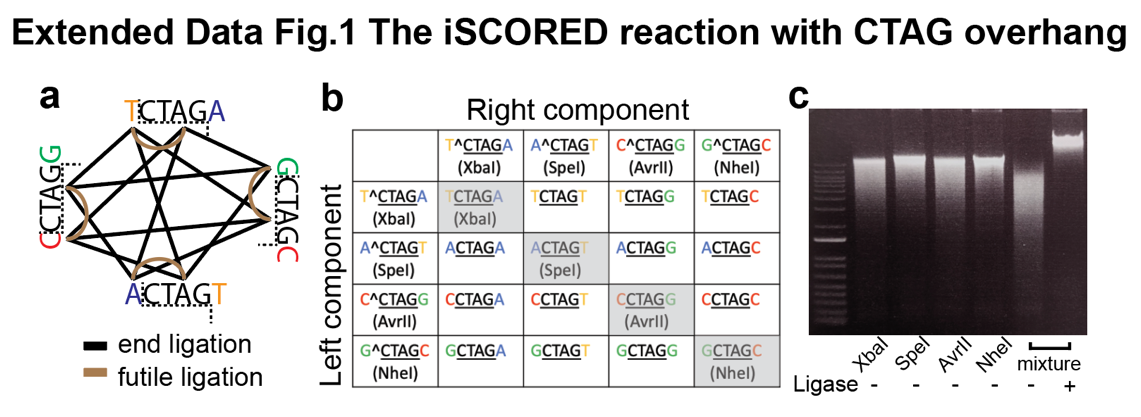

Figure S1: The iSCORED reaction with CTAG overhang as proof of principle. **a**, XbaI and SpeI recognize TCTAGA and ACTAGT, respectively. Upon enzymatic digestion, both release cohesive 5’CTAG overhangs, which could be re-ligated to TCTAGA, ACTAGT, TCTAGT, and ACTAGA with co-existing DNA ligase. While the first two products are susceptible to further digestion (by XbaI and SpeI, respectively, brown lines), TCTAGT and ACTAGA are irreversible final products (black lines). **b**, The likelihood of forming such irreversible ligations increases with the number of different restriction enzymes producing compatible ends (up to 75% for 6-mer REs releasing 4-base pair (bp) overhangs). **c**, Using control human genomic DNA (gDNA; NA12878 from Coriell Institute), the digested DNA units are reconstructed into larger fragments in the presence of T4 DNA ligase.

Figure S2

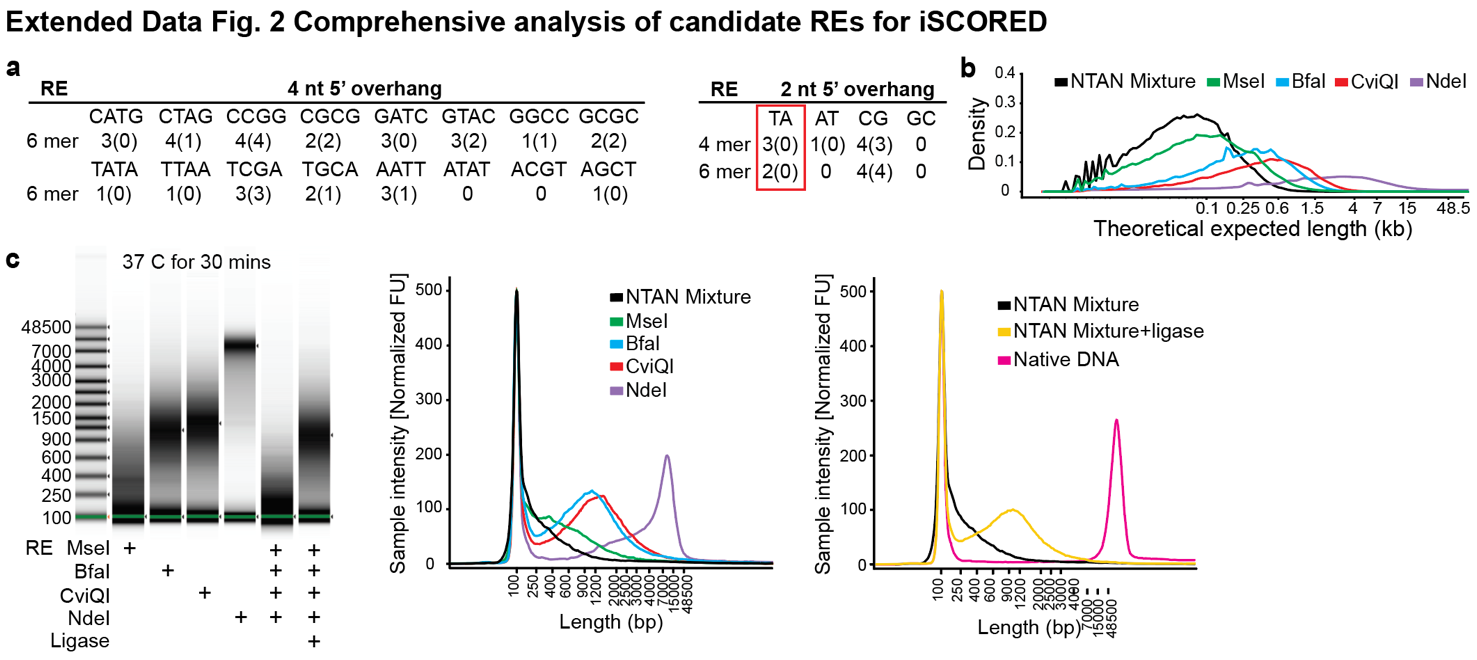

Figure S2: Comprehensive analysis of candidate REs for iSCORED. **a**, List of all enzymatic combinations for iSCORED (4 nt overhang in left panel and 2 nt overhang in right panel). The numbers denote available REs to produce such overhangs and numbers in the parentheses denote REs sensitive to CpG methylation. **b**, *In silico* estimation of NTAN cocktail mix using the complete human genome database as a reference (T2T-CHM13^40^). **c**, Quantitative electrophoresis (Agilent) to assess the digestion efficiencies of restriction endonuclease generating TA overhang and iSCORED reaction.

Figure S3

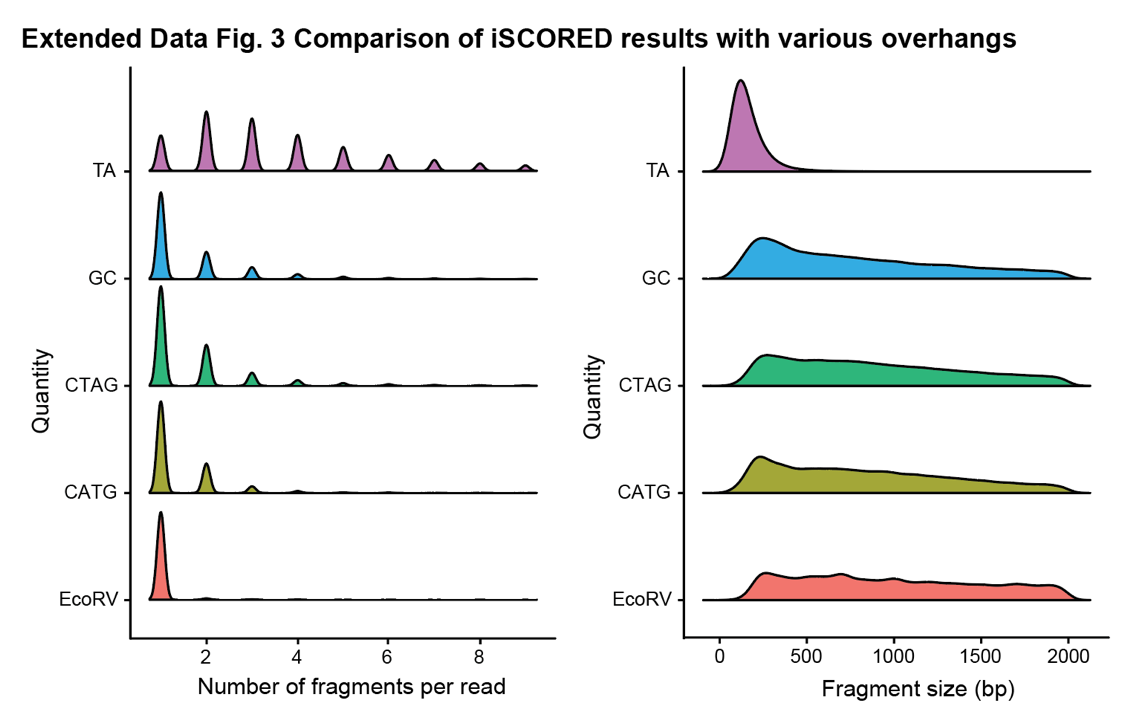

Figure S3: Comparison of iSCORED results with various overhangs. Distribution of concatenation numbers and sequenced lengths in iSCORED reactions utilizing cocktail combinations generating 2-nt and 4-nt 5’ overhangs.

Figure S4

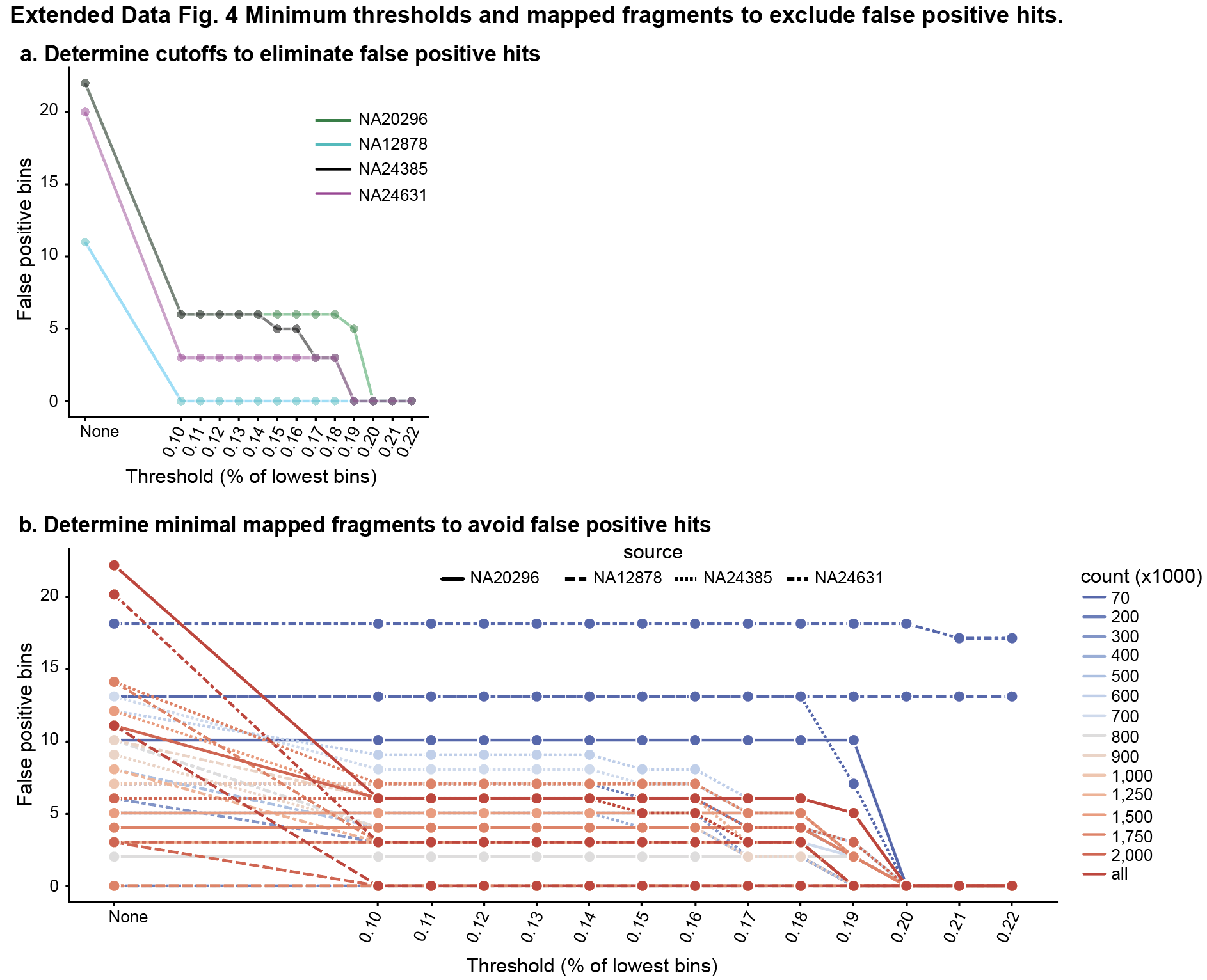

Figure S4: Minimum thresholds and mapped fragments to exclude false positive hits. **a**, False positive hits were eliminated from the full datasets of control gDNA samples by setting a minimum threshold of 0.2 percentile. **b**, The mapped fragments count of 200k or higher, in conjunction with the 0.2 percentile threshold is sufficient to eliminate all false positive hits.

Figure S5

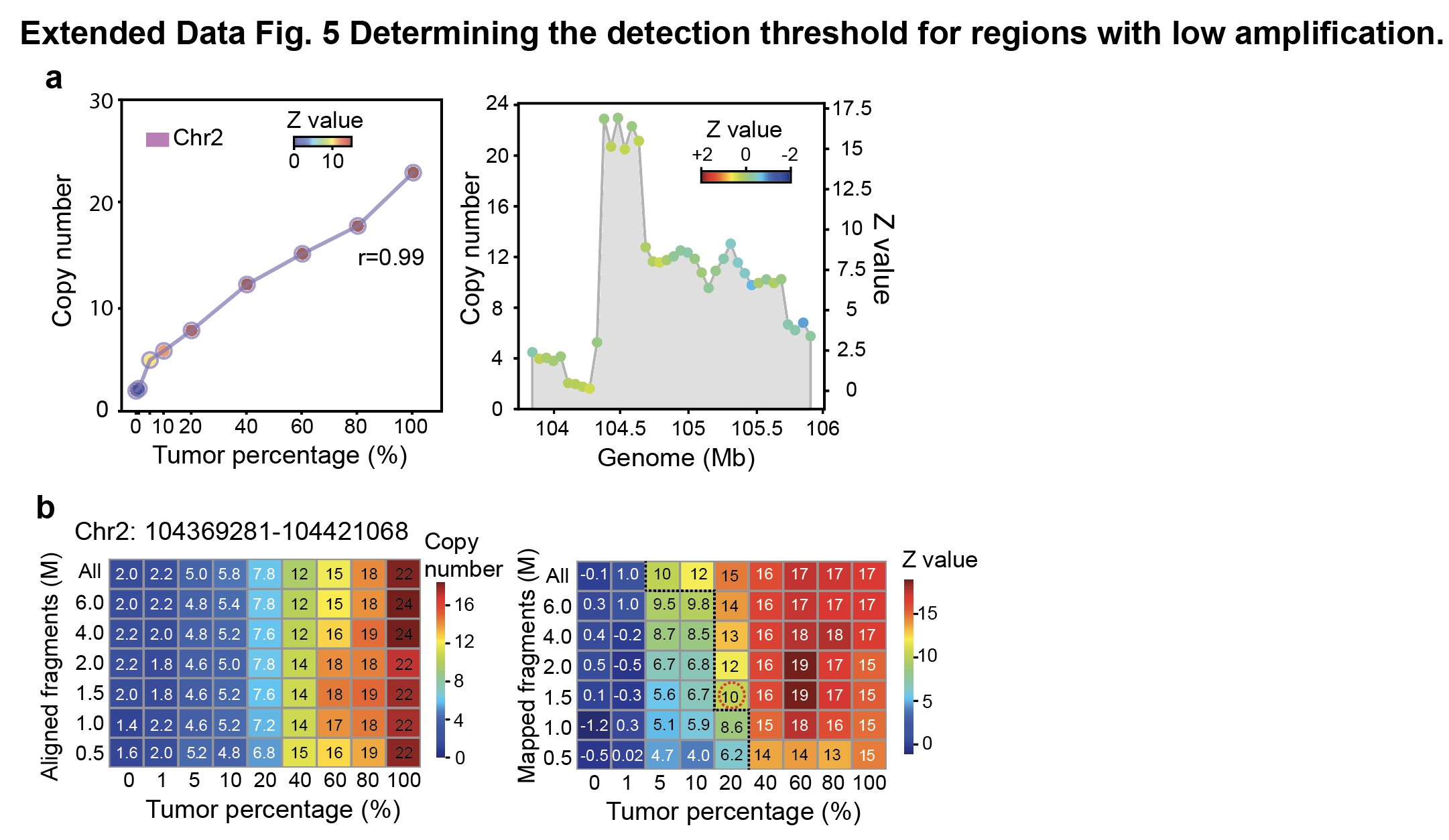

Figure S5: Determining the detection threshold for regions with low amplification. Focal gene amplification (22 copies) was detected in chromosome 2 of an adenosquamous carcinoma. We evaluated the sensitivity of our approach in two dimensions: the fragment counts and the tumor percentages. Using a Z-score cutoff of 10, we were able to reliably detect the amplification in samples with 20% tumor purity, based on a dataset of 1.5 million mapped fragments. Pearson correlation of the data is shown.

Figure S6

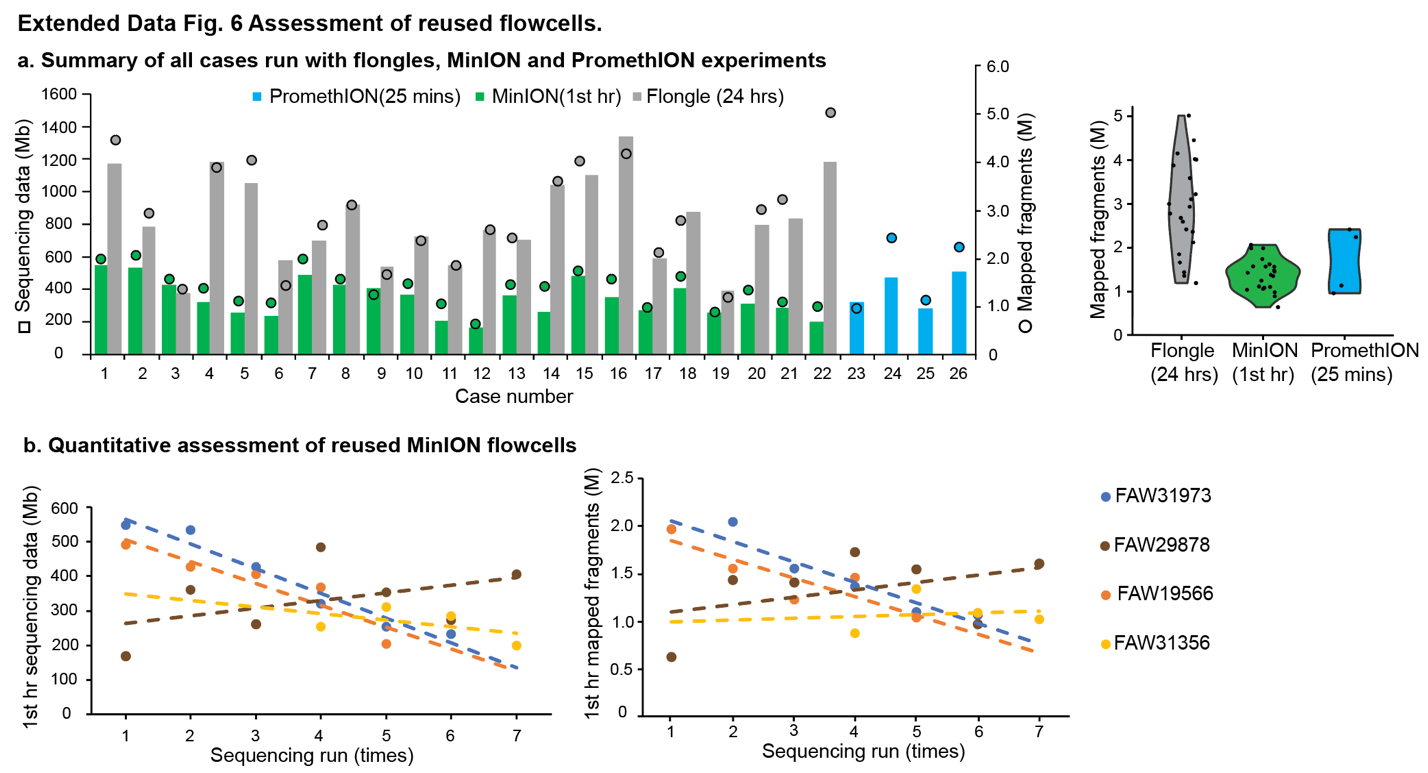

Figure S6: Assessment of reused flowcells. **a**, Total sequenced data (Mb) and mapped fragments from MinION and flongle runs. The usable MinION flowcells generate [(1.38 ± 0.08) x 10^6^ (SEM)] mapped fragments within one hour whereas flongles generate[(2.9 ± 0.23) x 10^6^ (SEM)] mapped fragments during 24 hours of sequencing. **b**, Exact sequencing data and mapped fragments of all samples in the reused MinION flowcells.

Figure S7

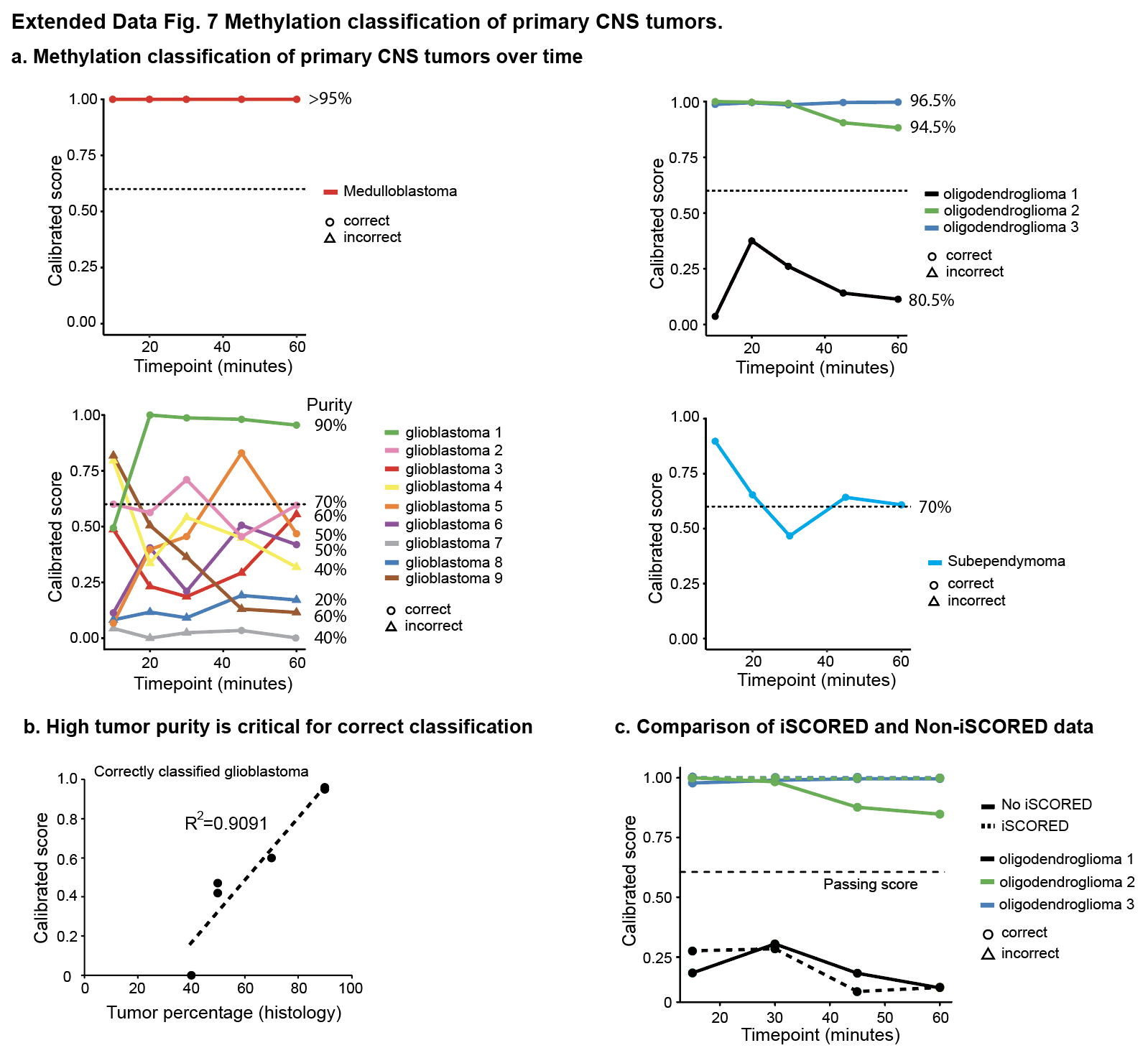

Figure S7: Methylation classification of primary CNS tumors. **a**, Calibrated scores across various intervals from the initiation of sequencing. **b**, Calibrated scores of the six correctly classified glioblastomas and corresponding tumor percentages by histological estimation. **c**, Comparison of methylation classification with intact gDNA (non-iSCORED) and iSCORED datasets with three oligodendroglioma cases.

Figure S8

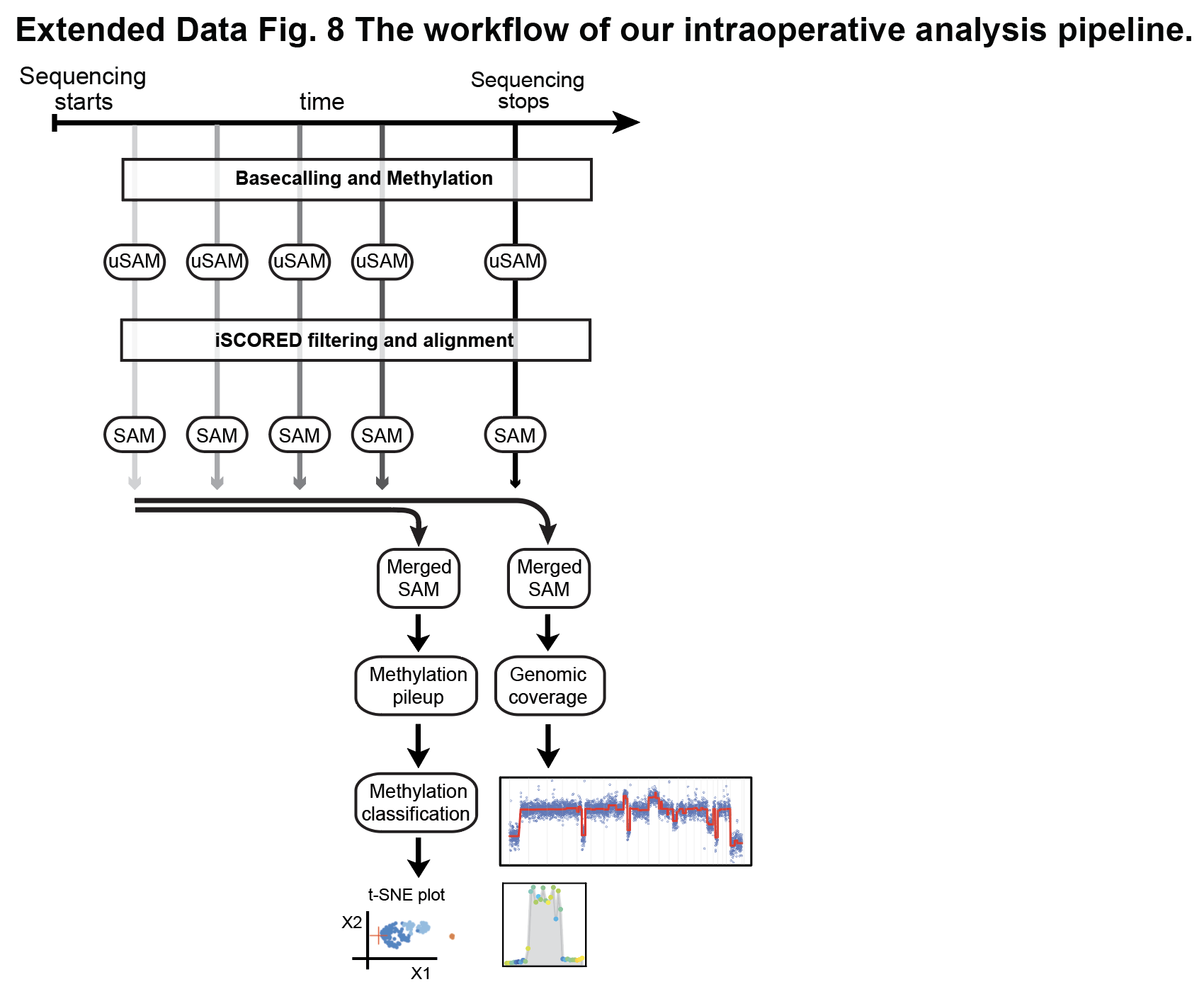

Figure S8: The workflow of our intraoperative analysis pipeline. Before beginning to sequence a configuration file is modified to the appropriate run parameters. Once sequencing begins the analysis pipeline is initiated. It periodically gathers the new fast5 files, basecalls them, extracting modification information, filters them and aligns them with the appropriate parameters for methylation and CNV/amplification analysis. At 40 minutes the aligned files for methylation are merged, the methylation call pileup is produced and used to classify the methylation class of the sample. At 55 minutes the aligned files for the CNV/amplification analysis are merged, one read from each duplex pair is removed, and the fragments per bin are calculated and used in the generation of a CNV plot and amplification analysis.

Figure S9: Comprehensive genome wide CNVs and oncogene promoter methylation analysis of the CNS tumor in the cohort

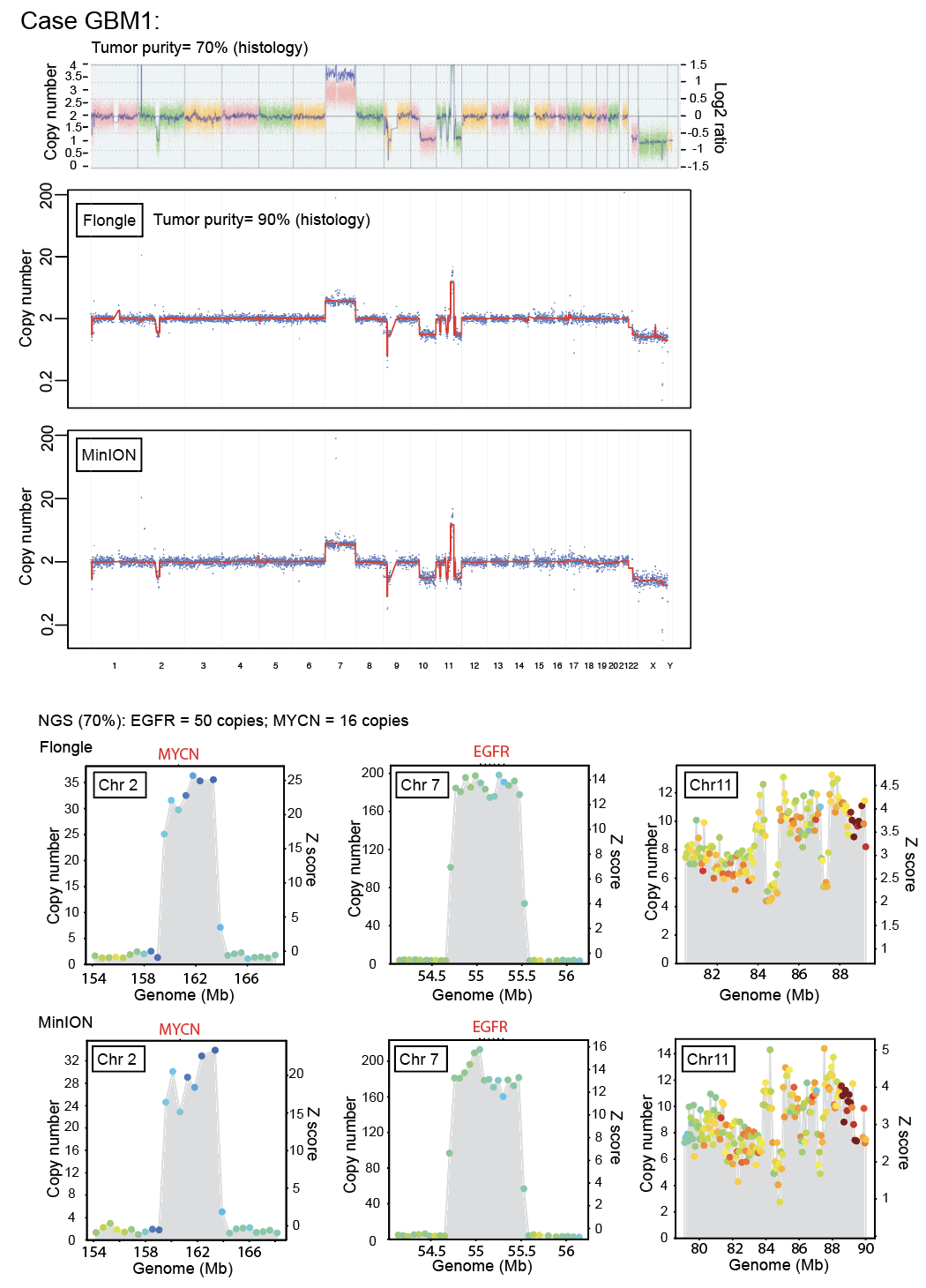

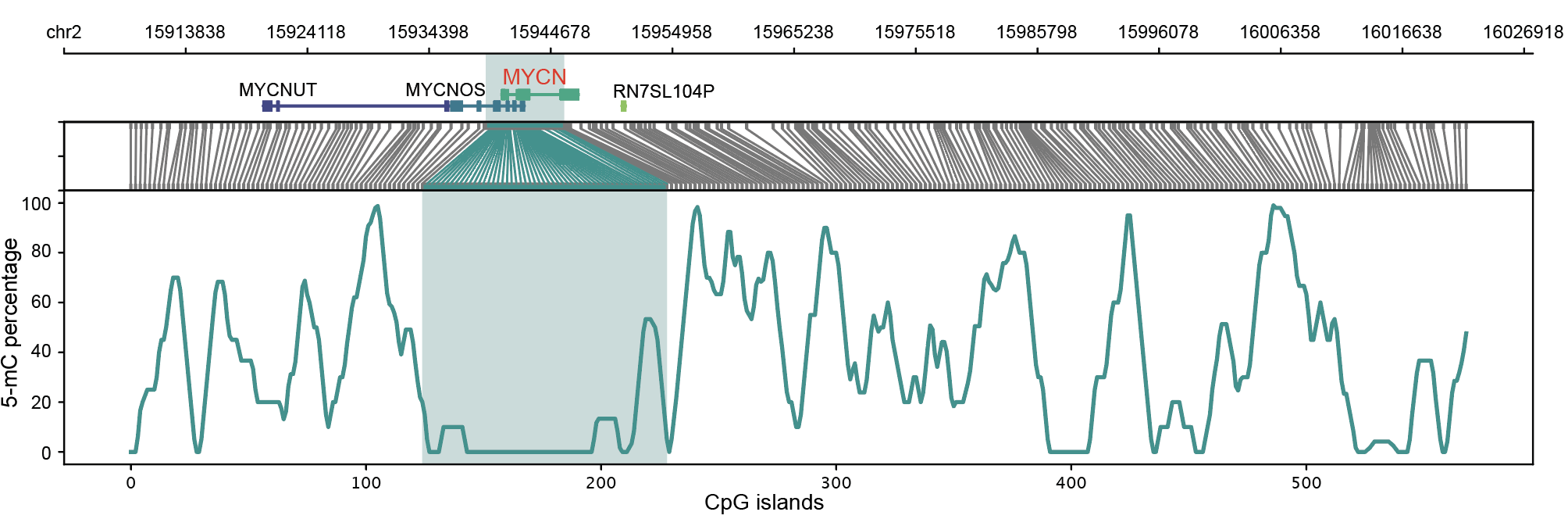

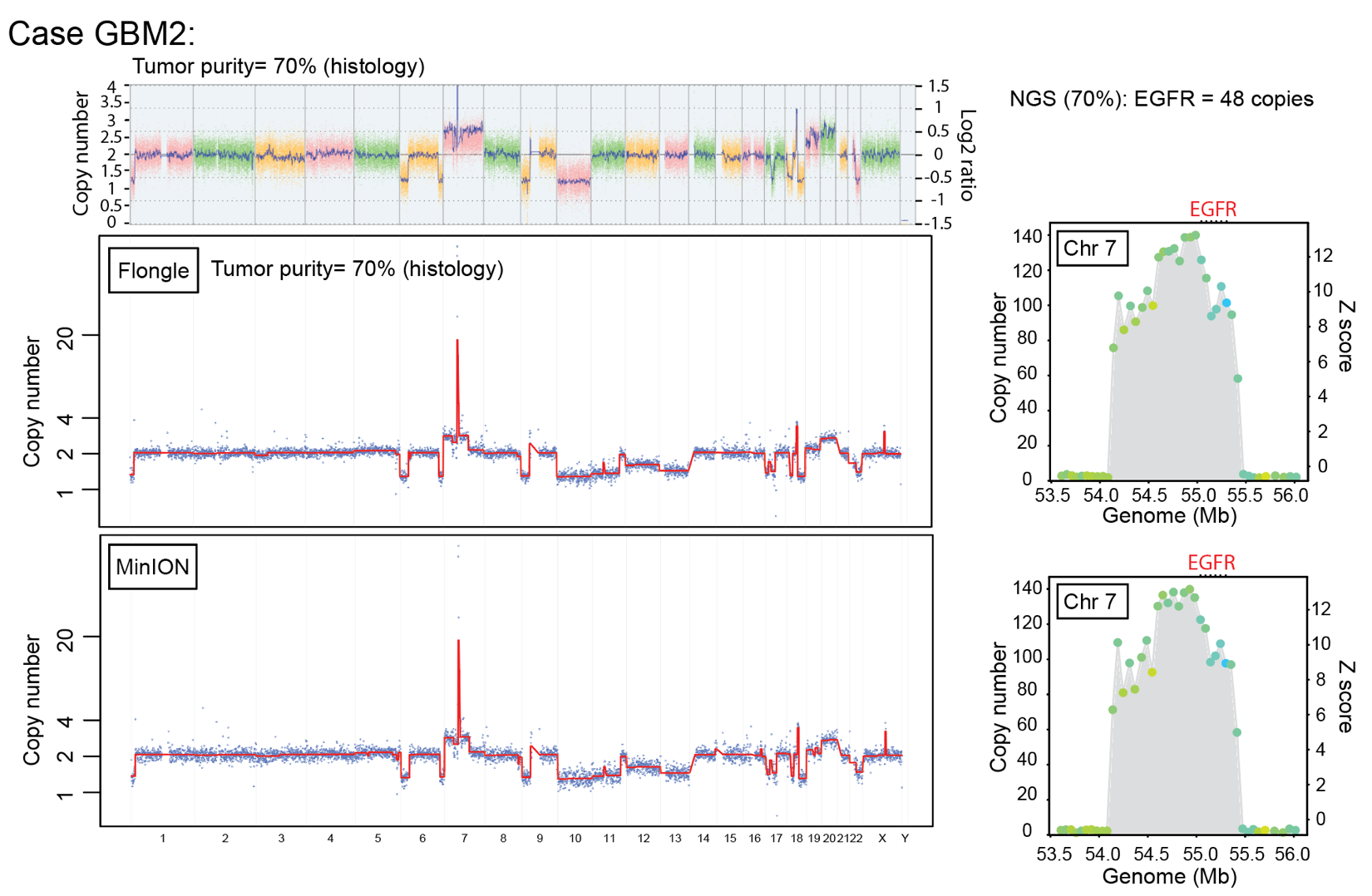

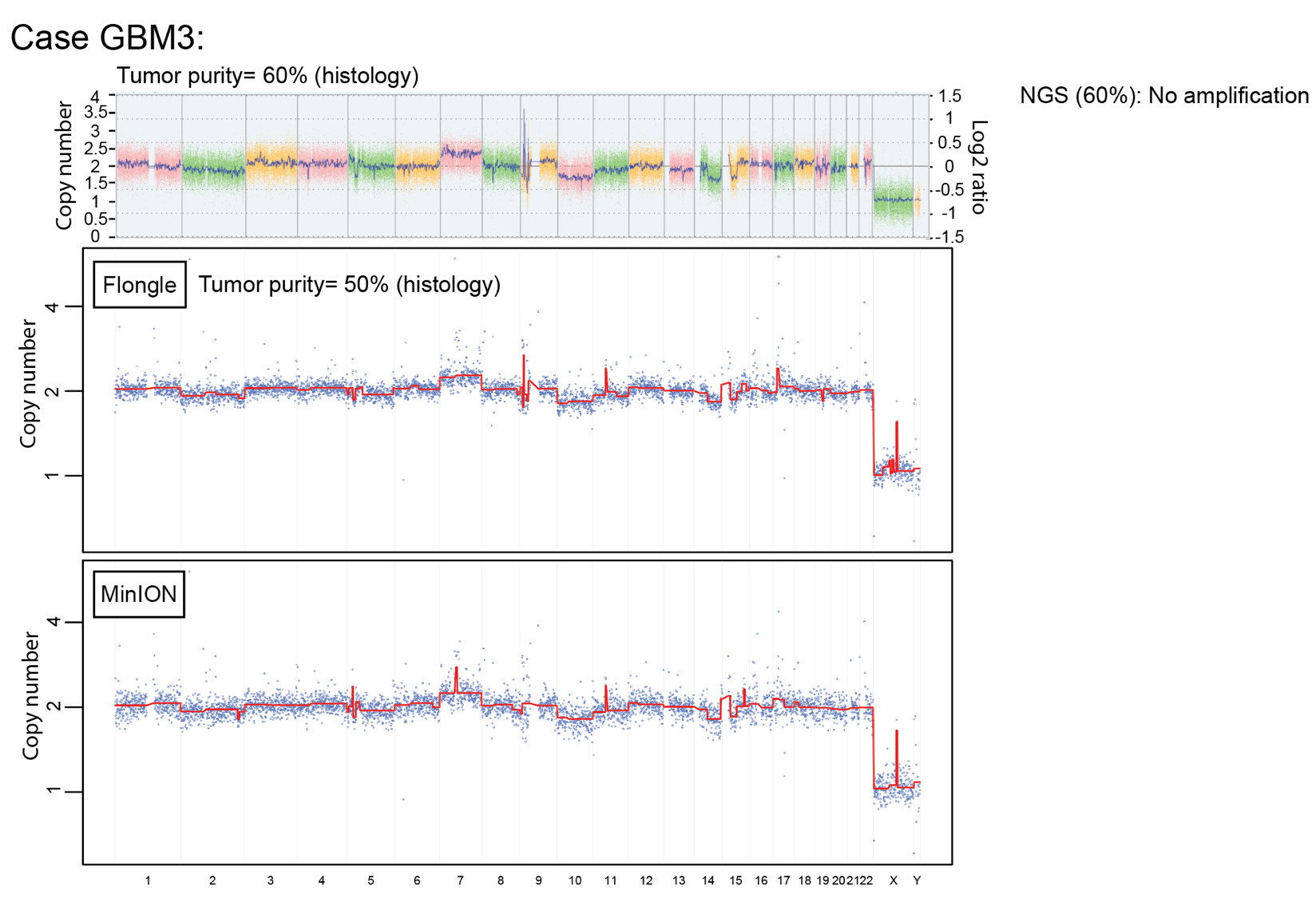

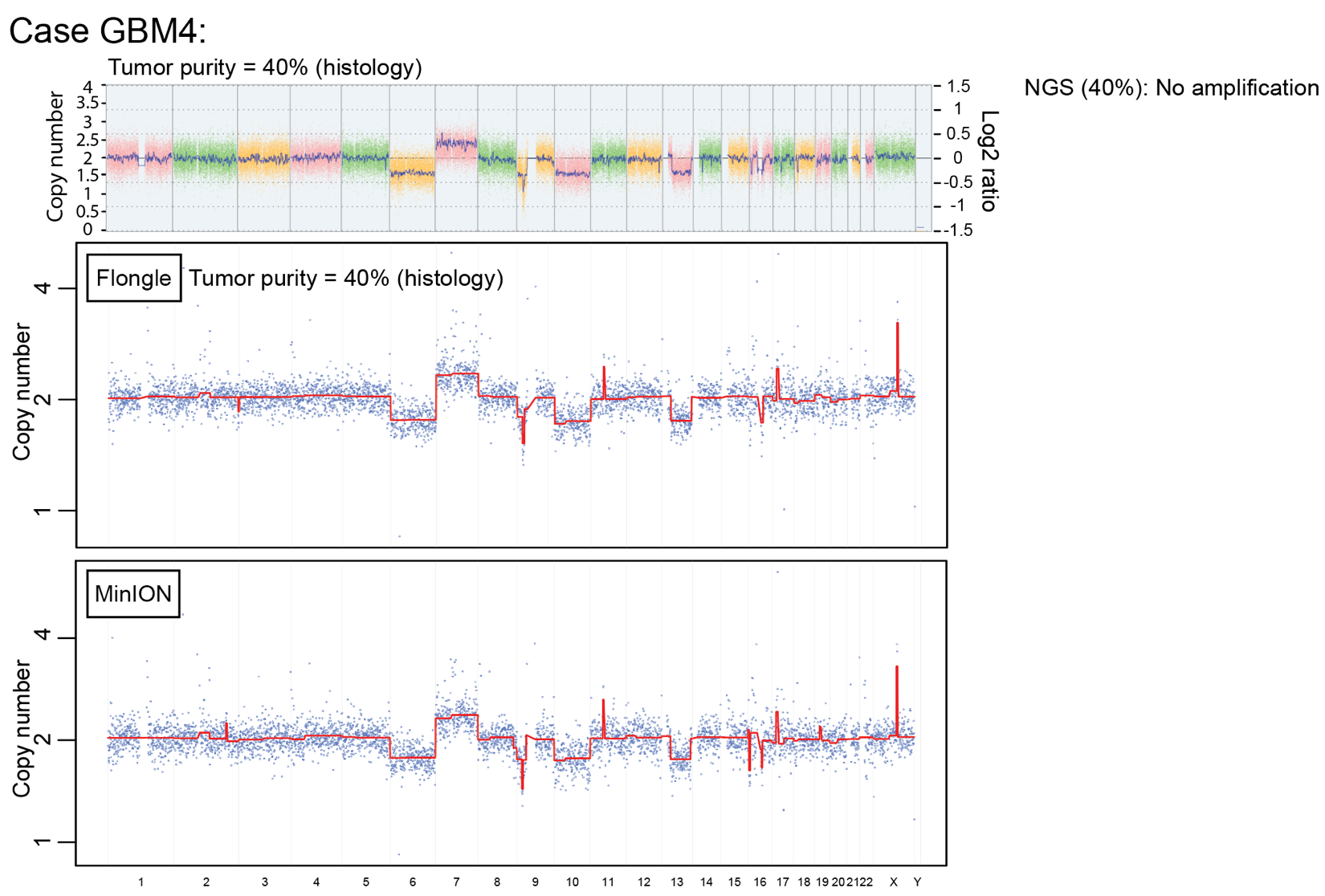

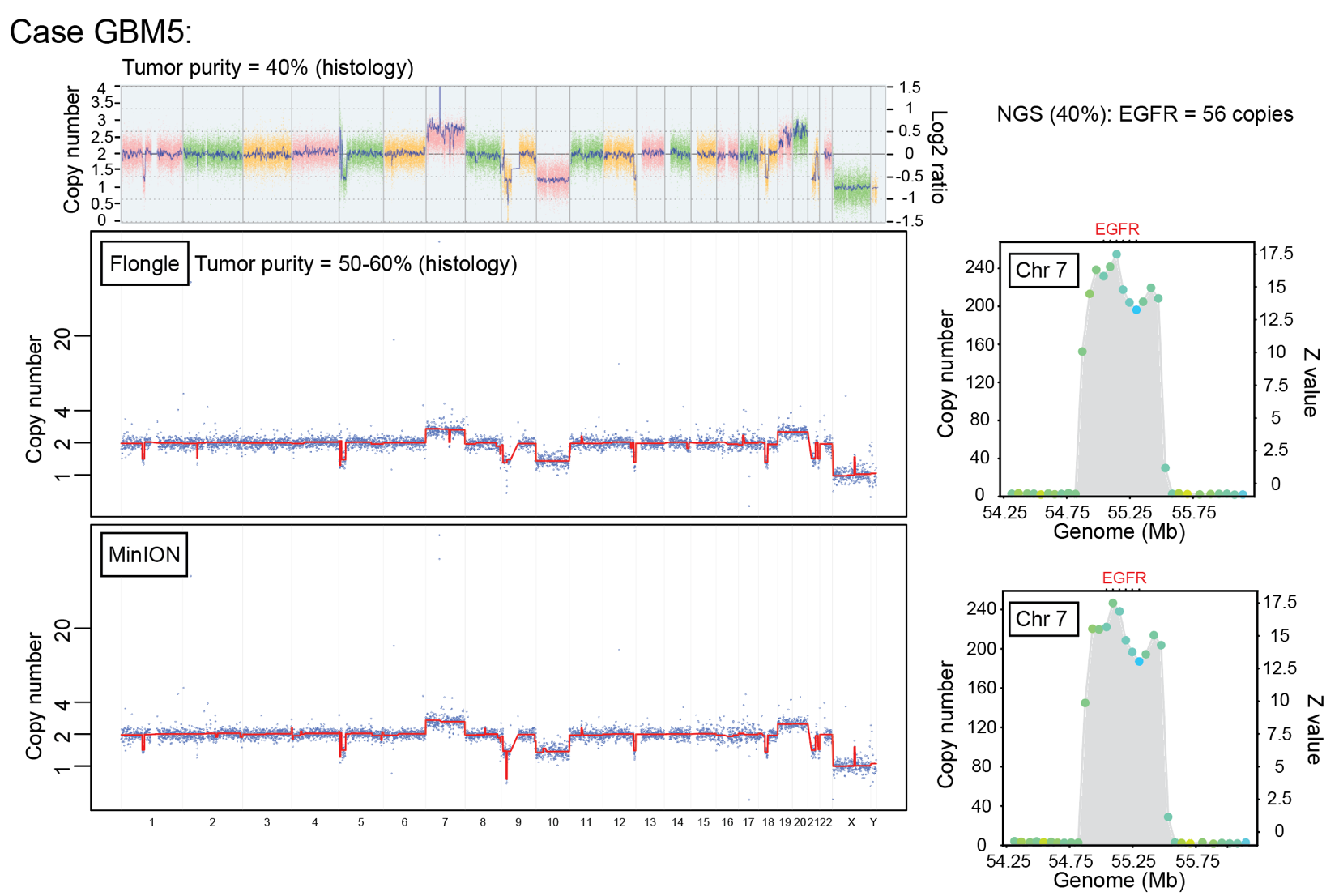

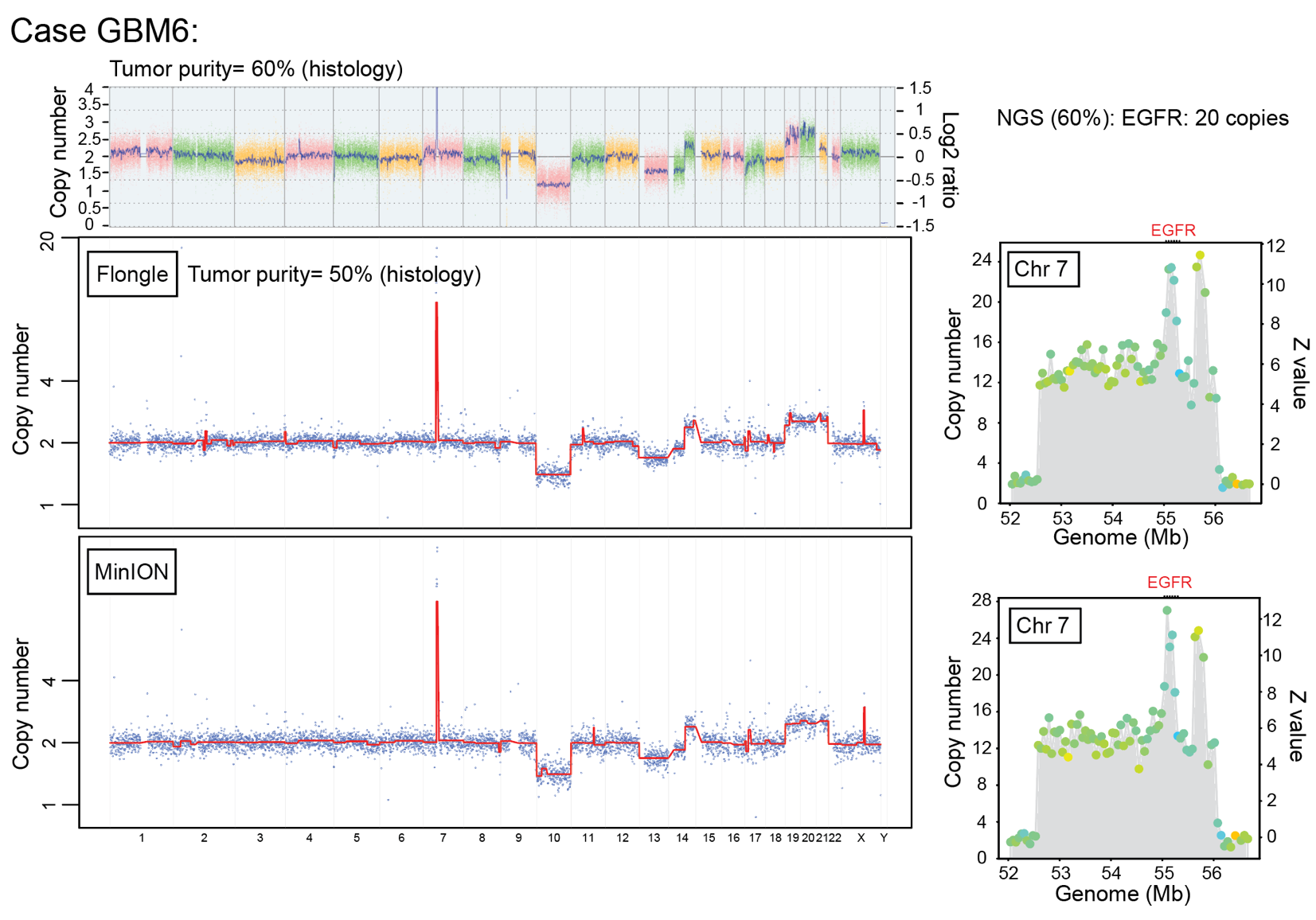

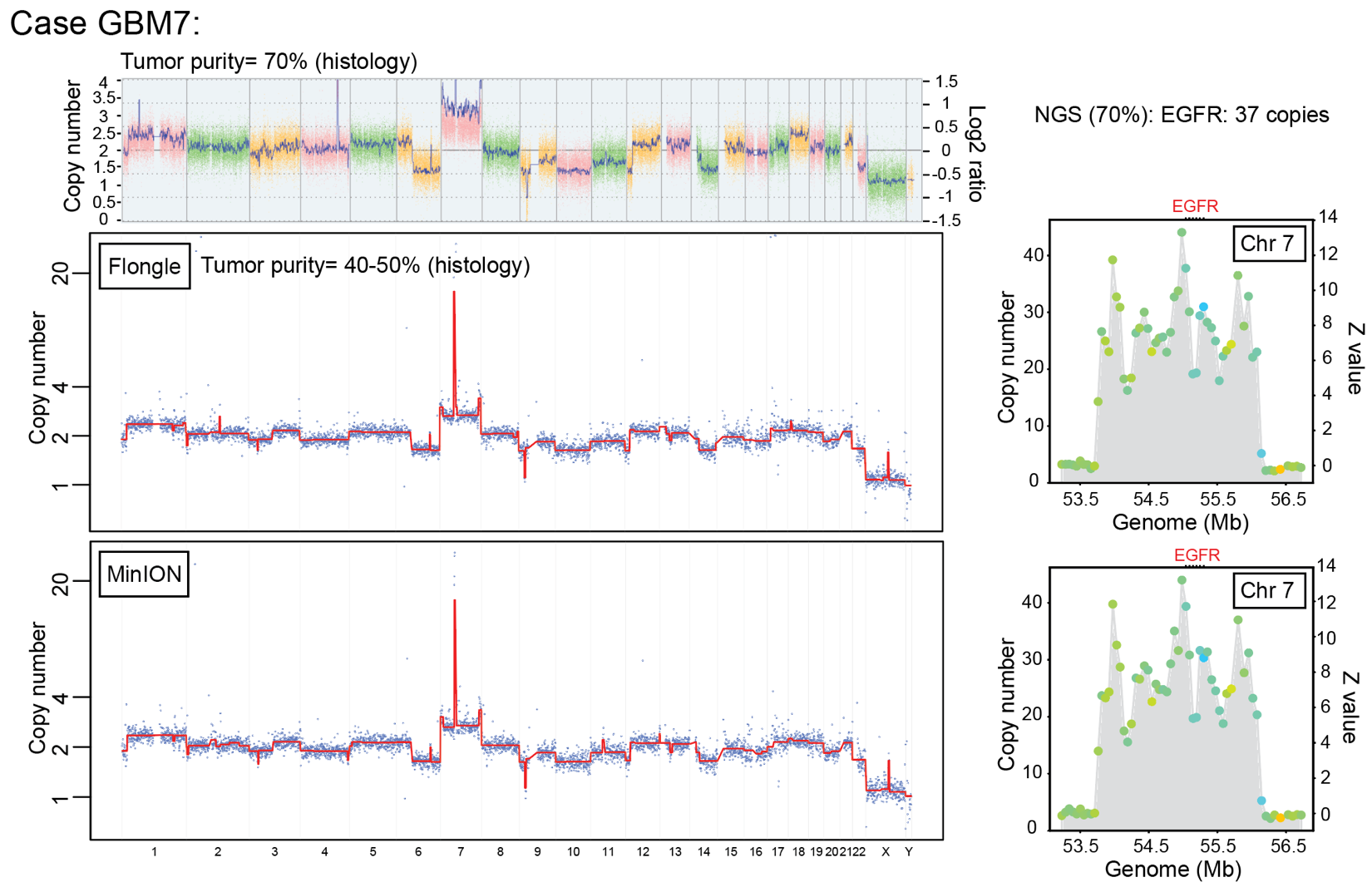

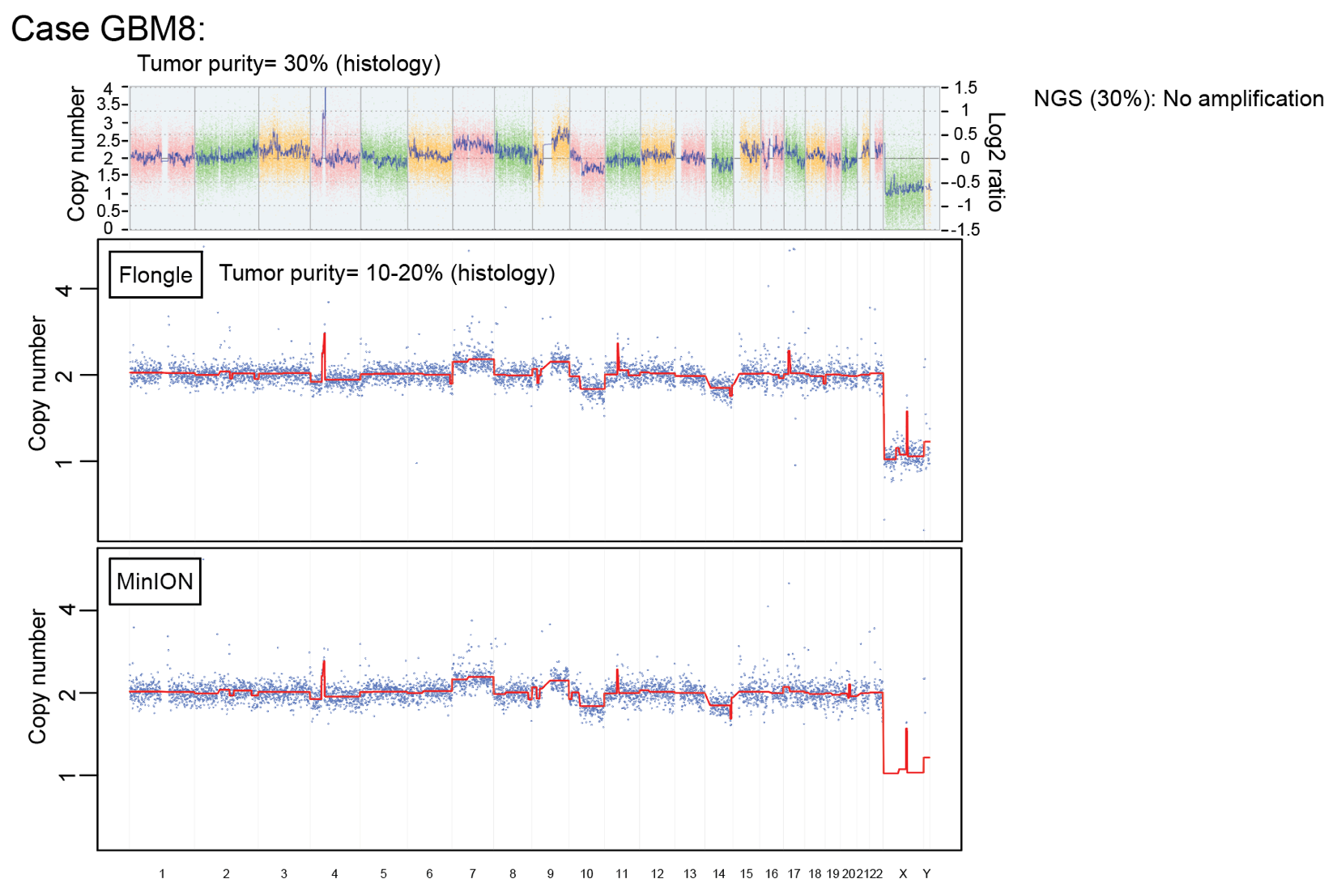

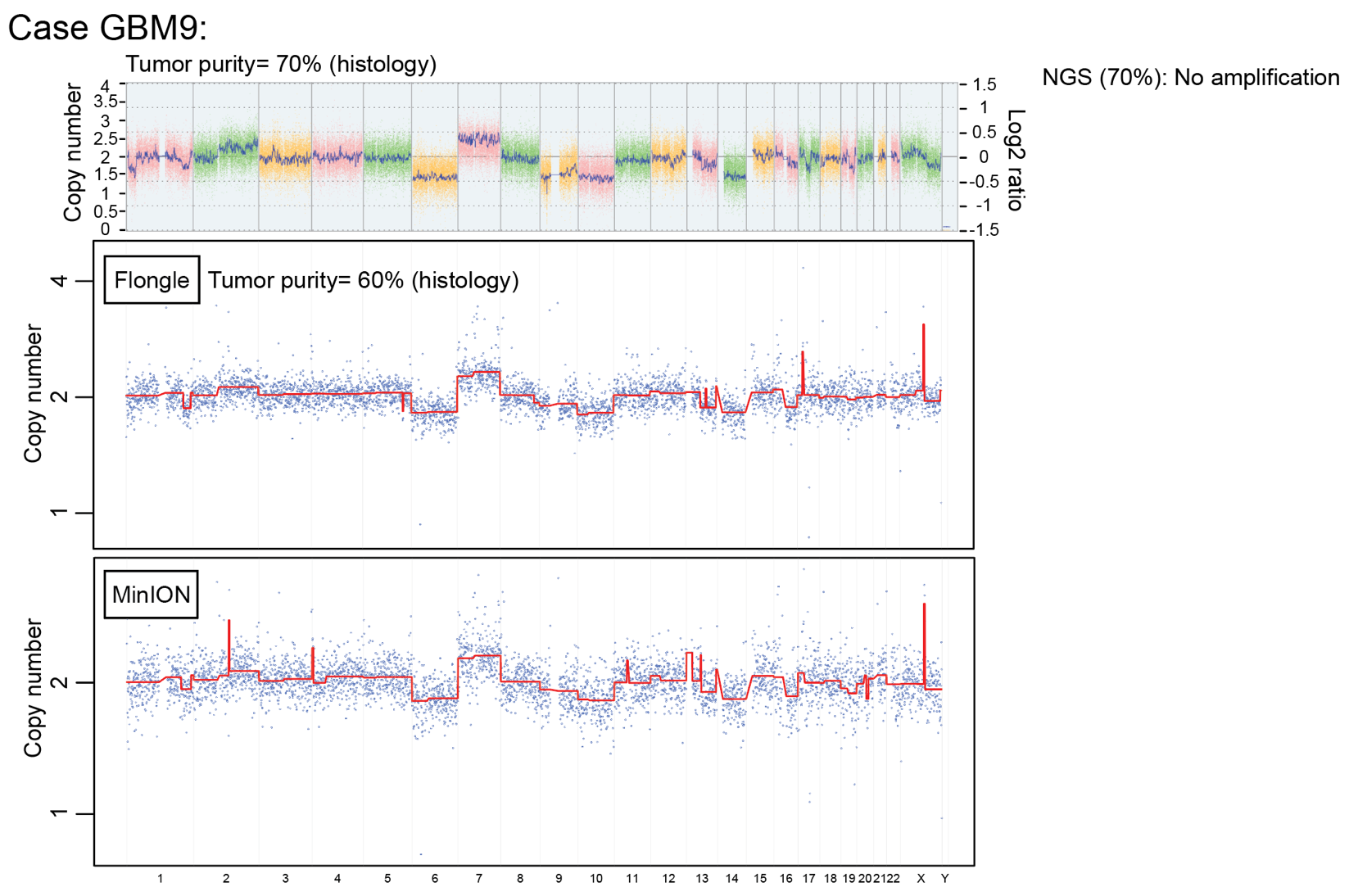

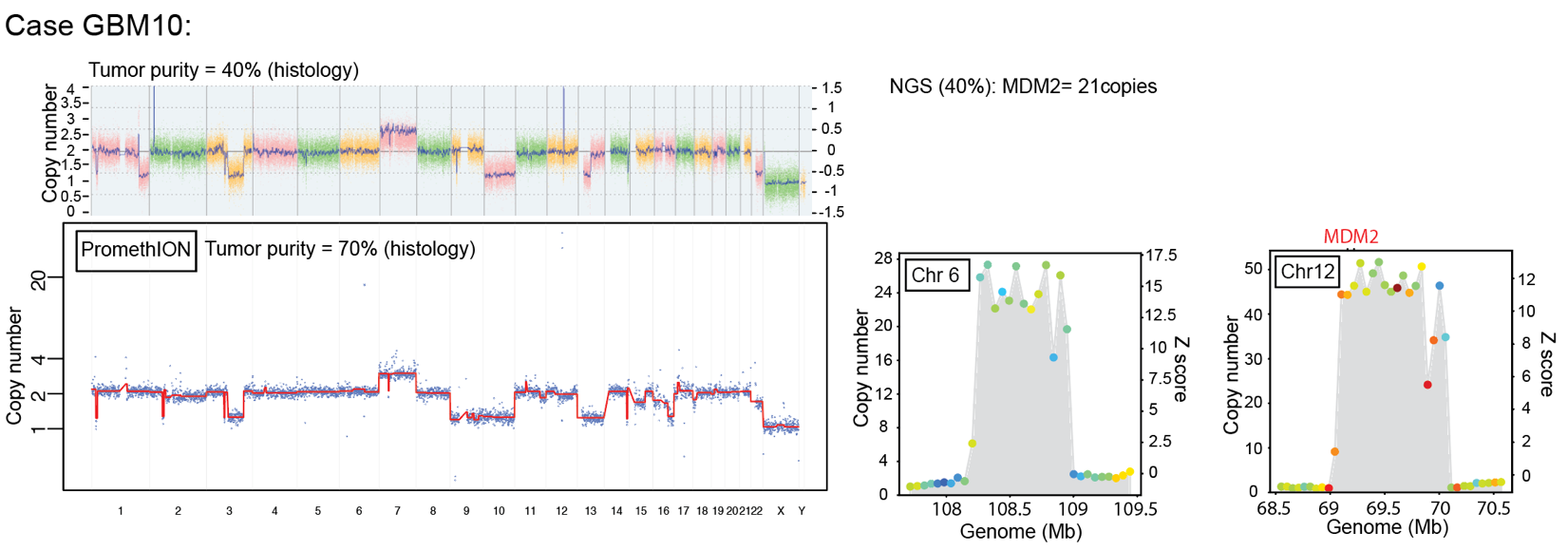

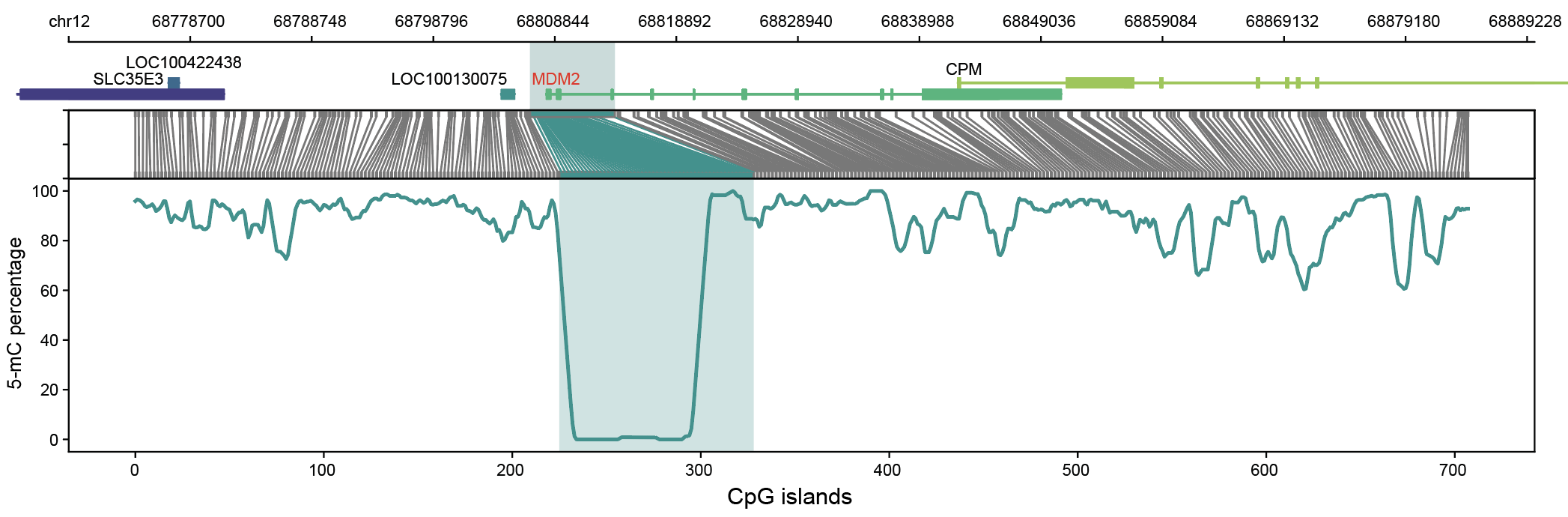

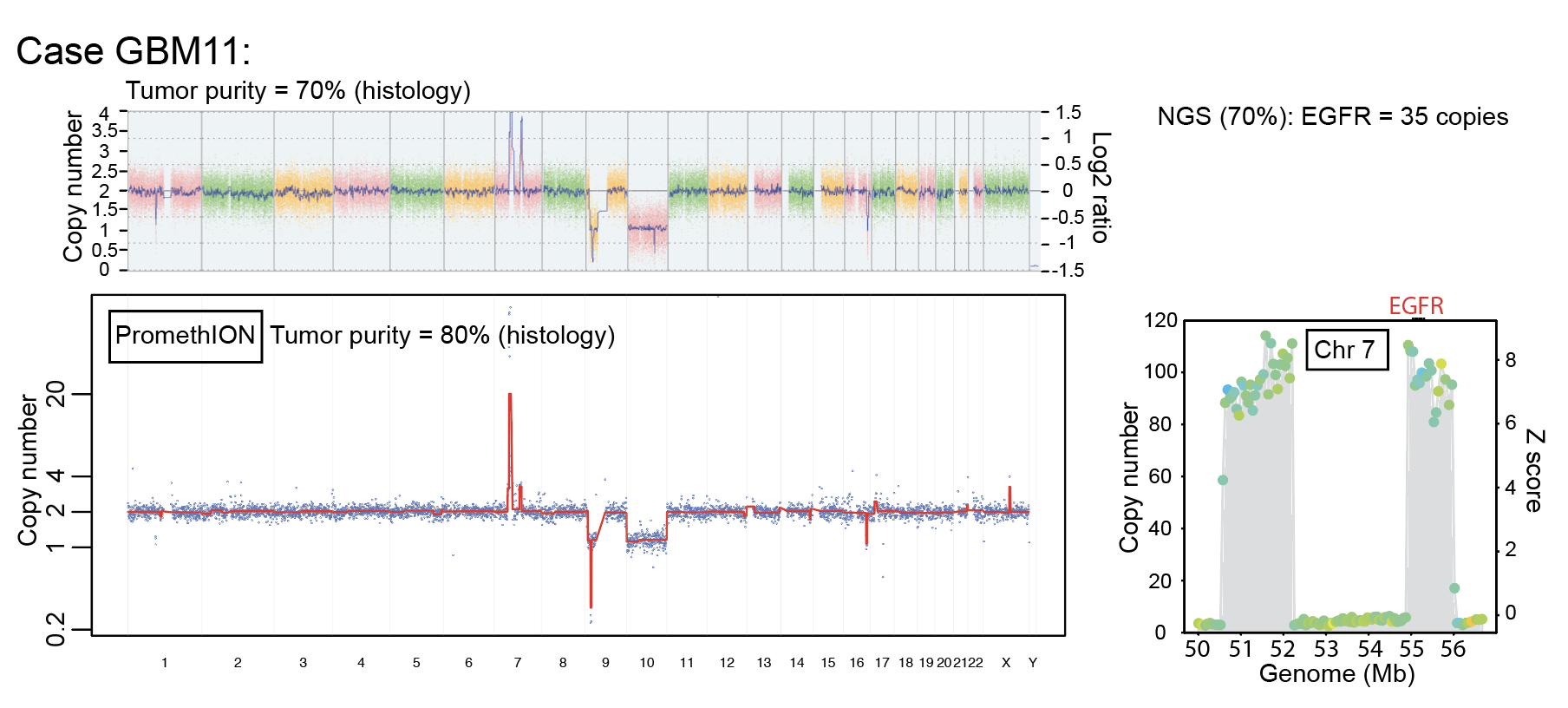

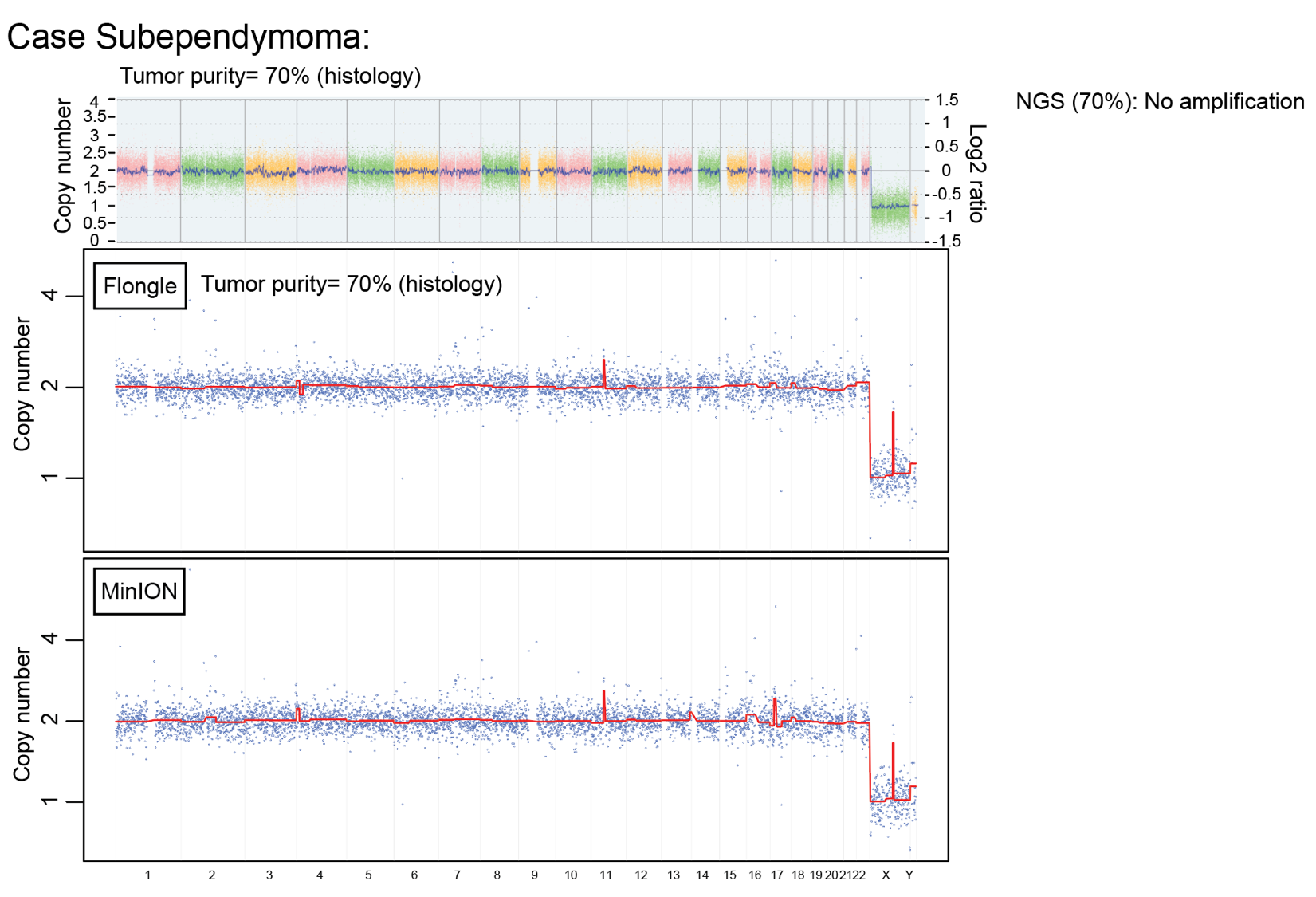

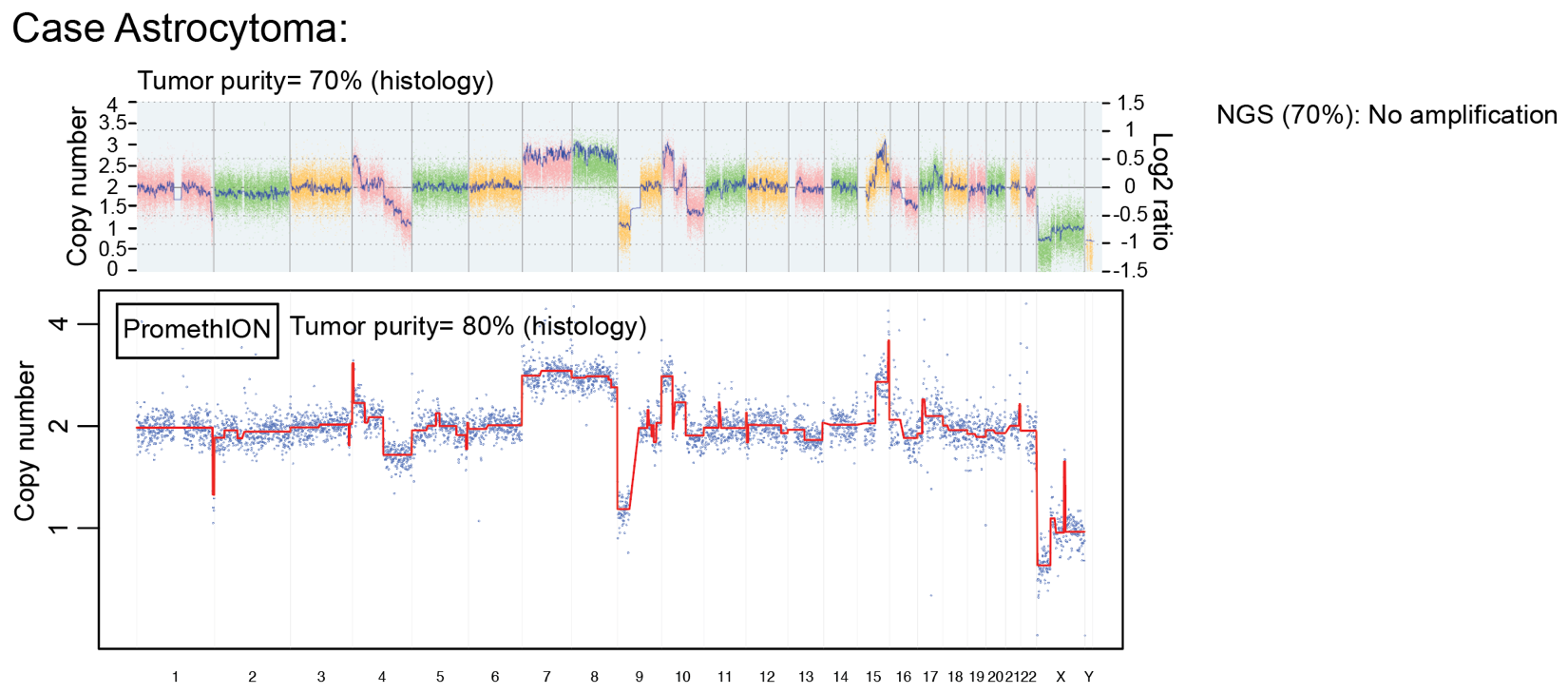

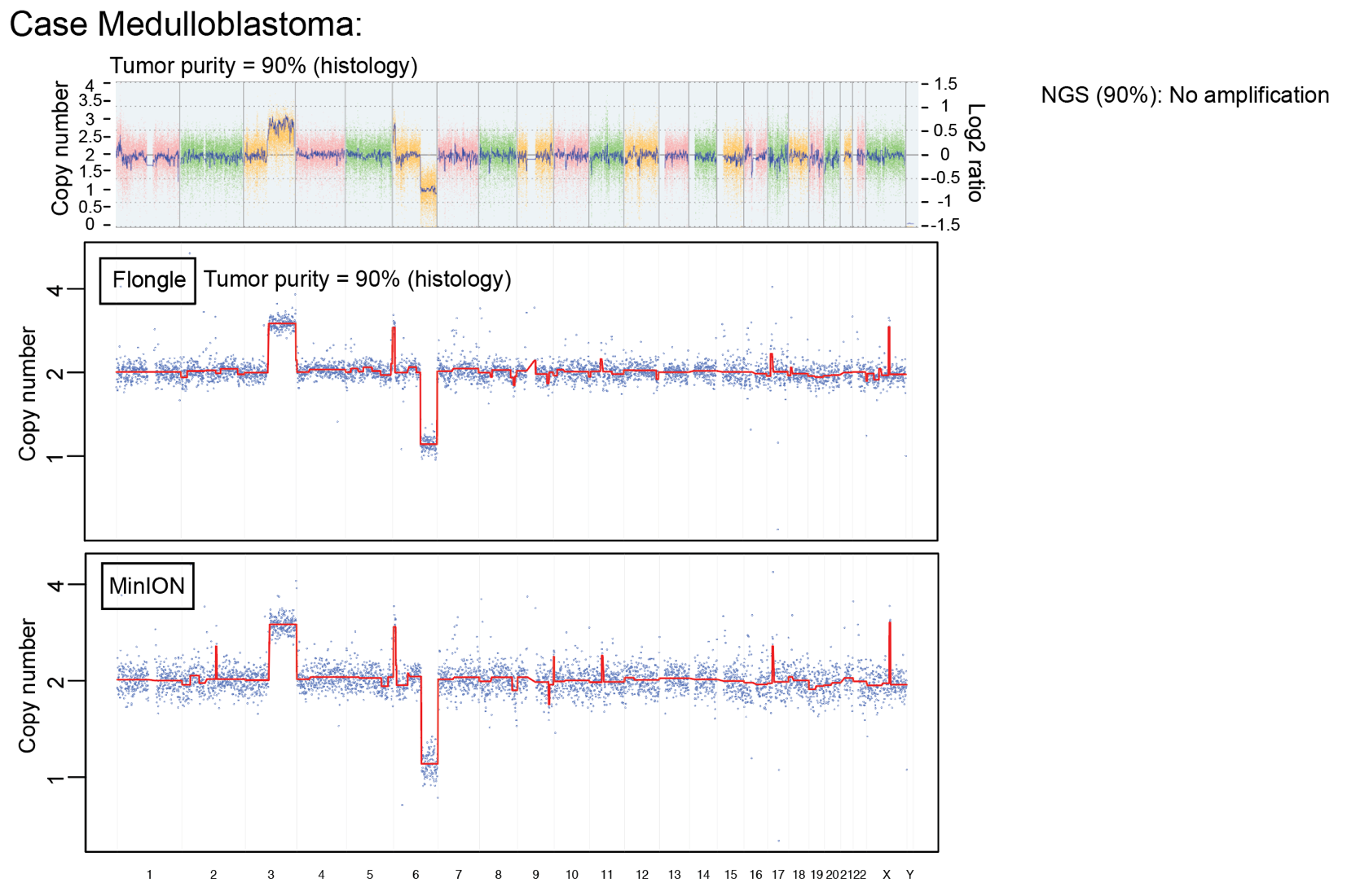

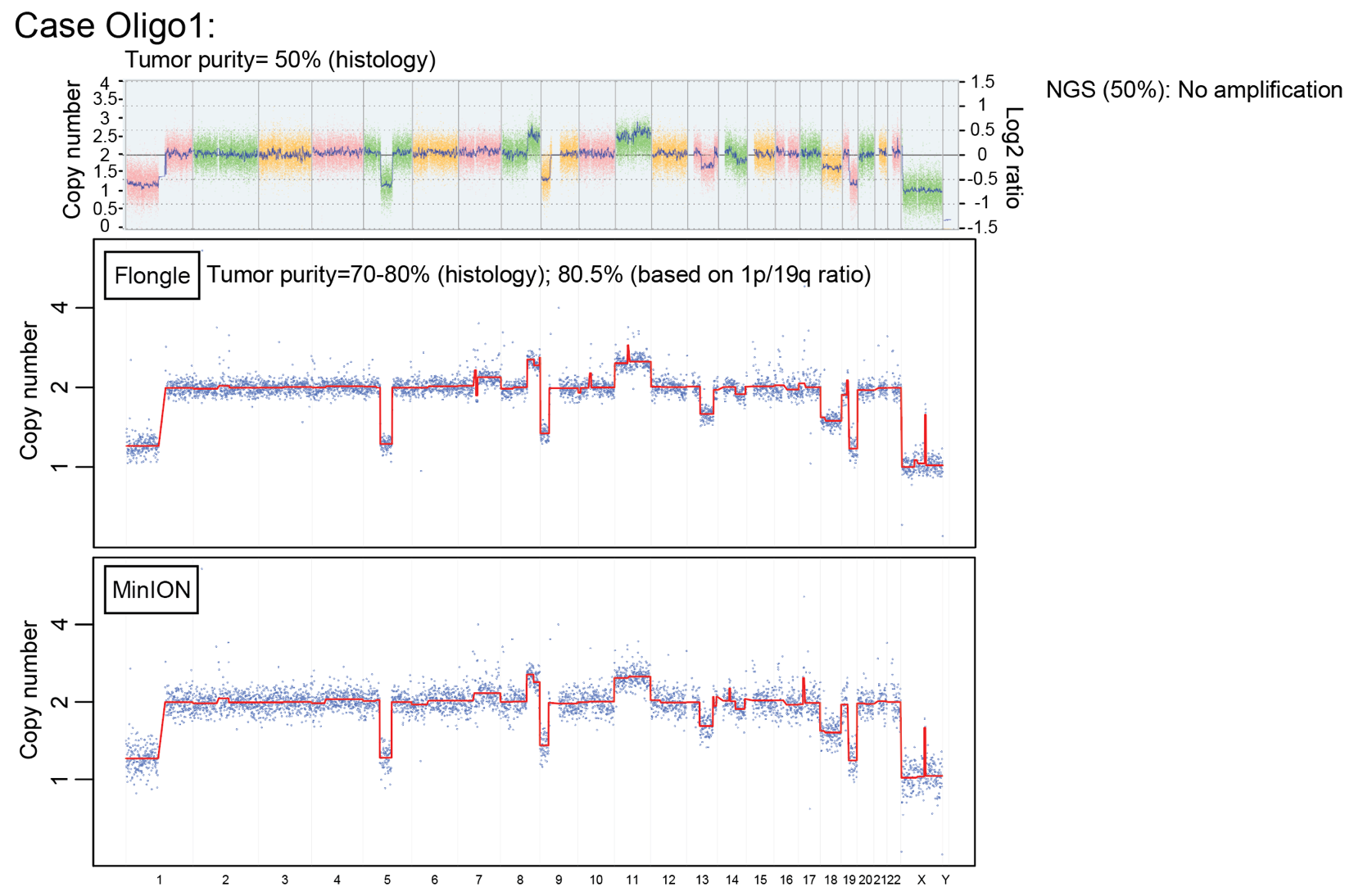

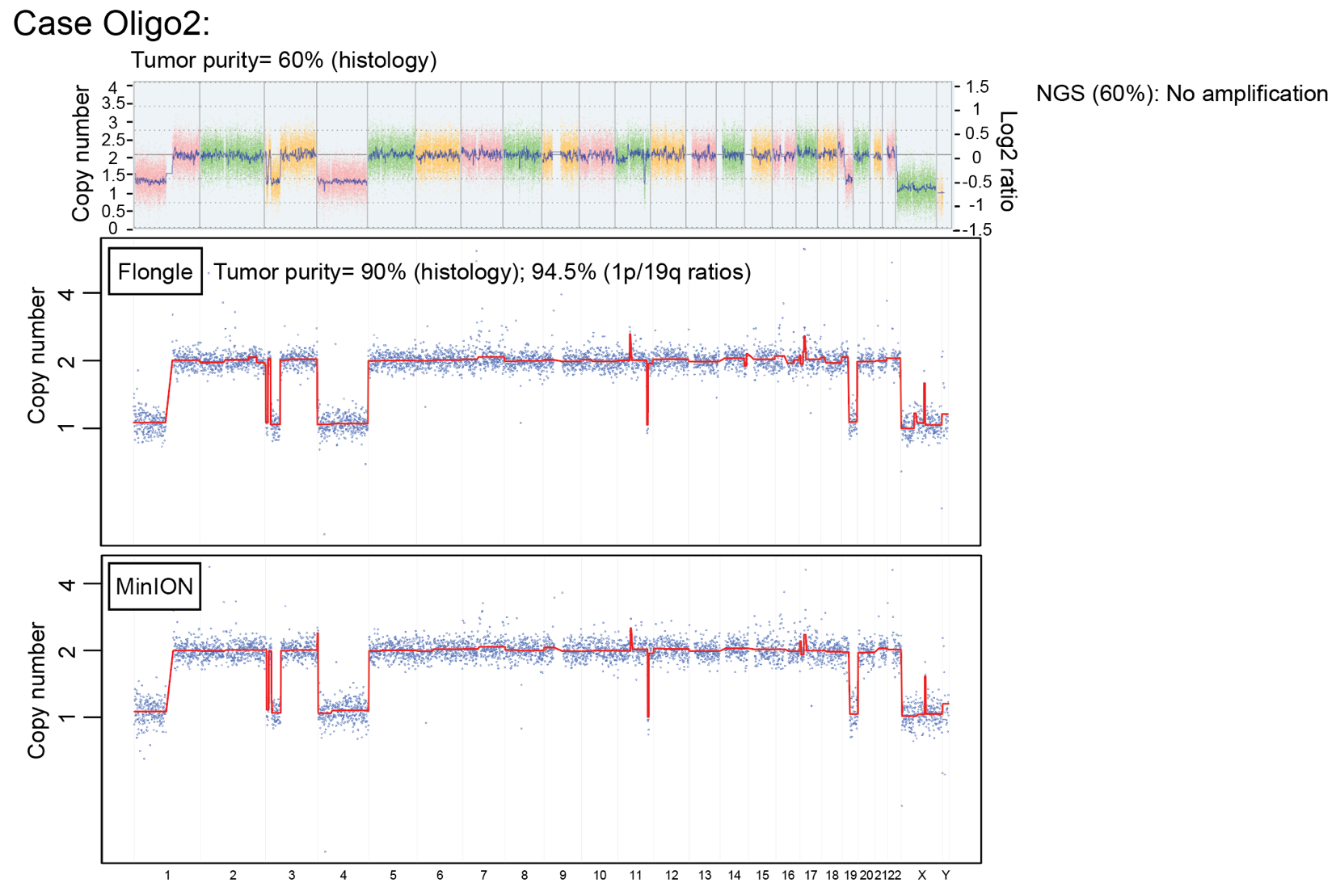

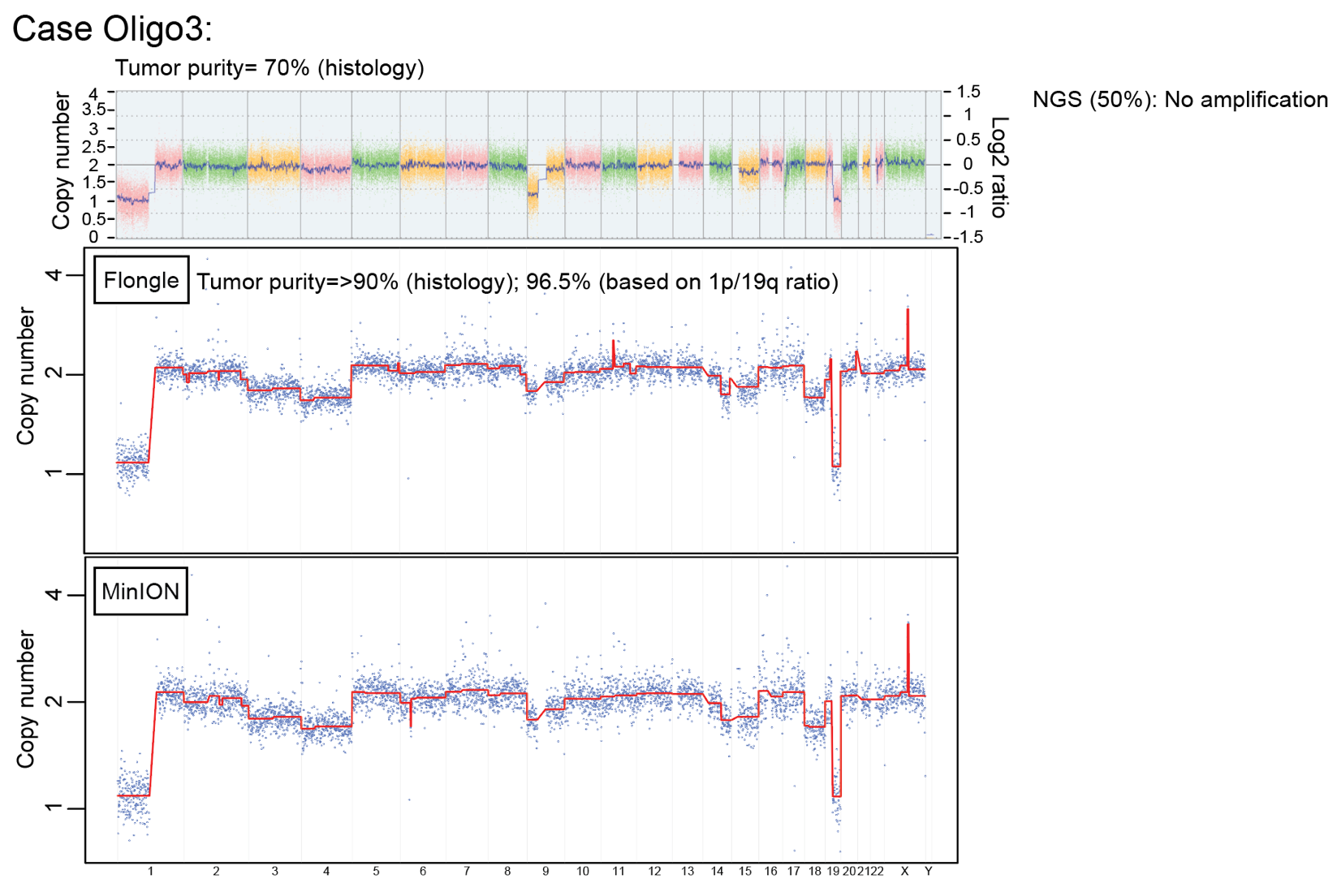

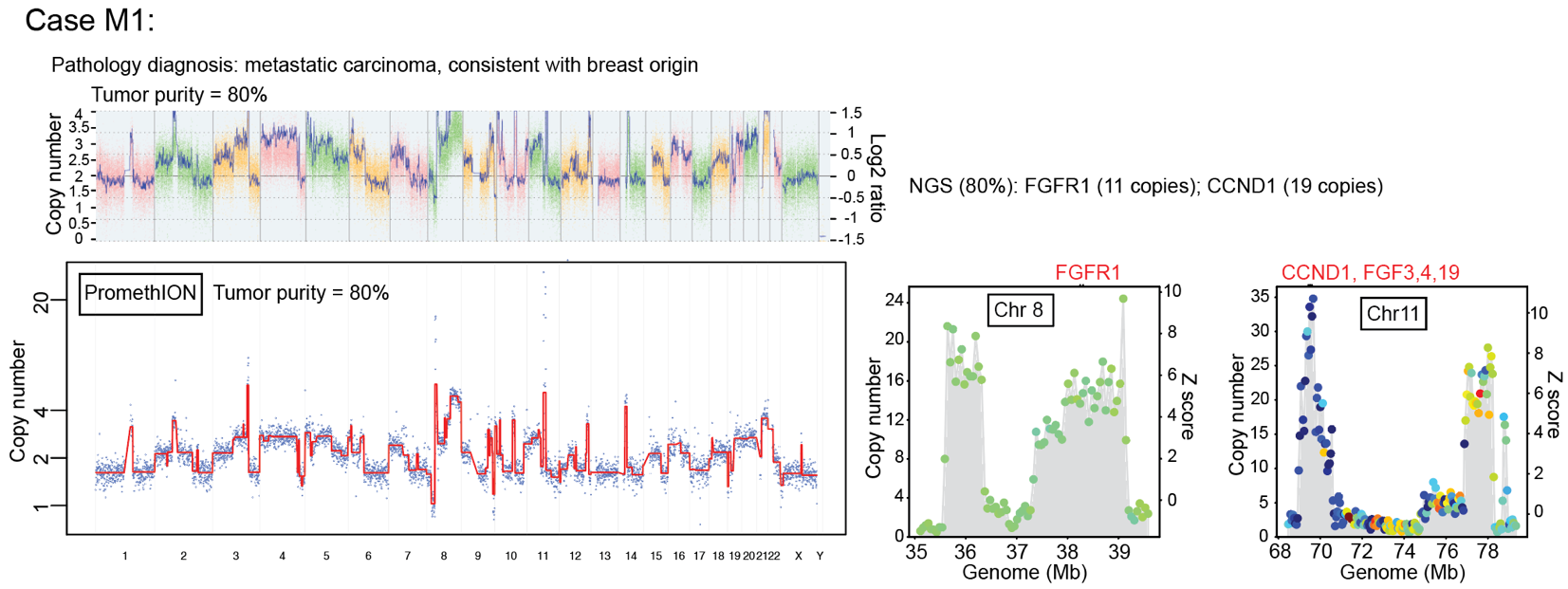

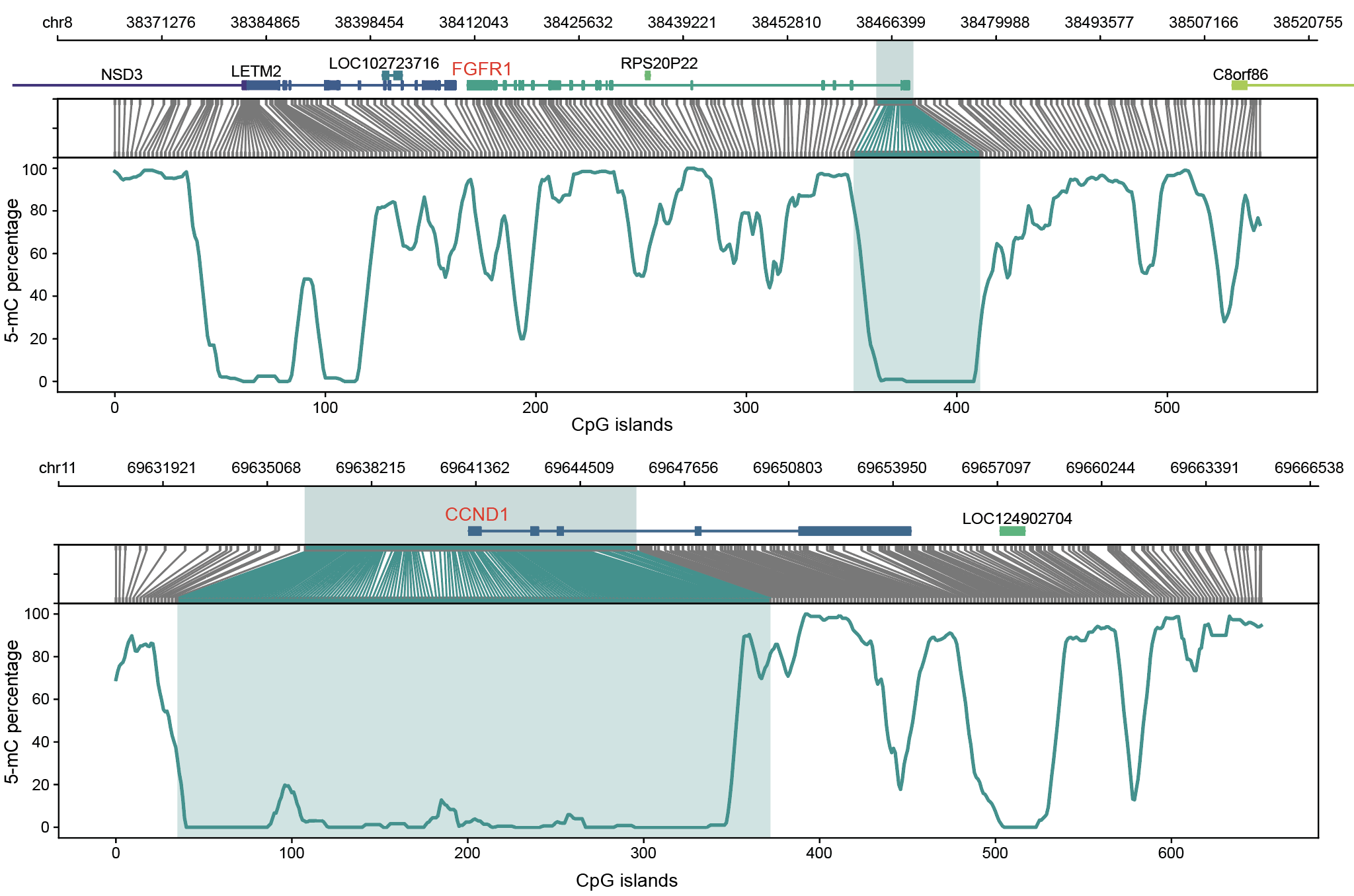

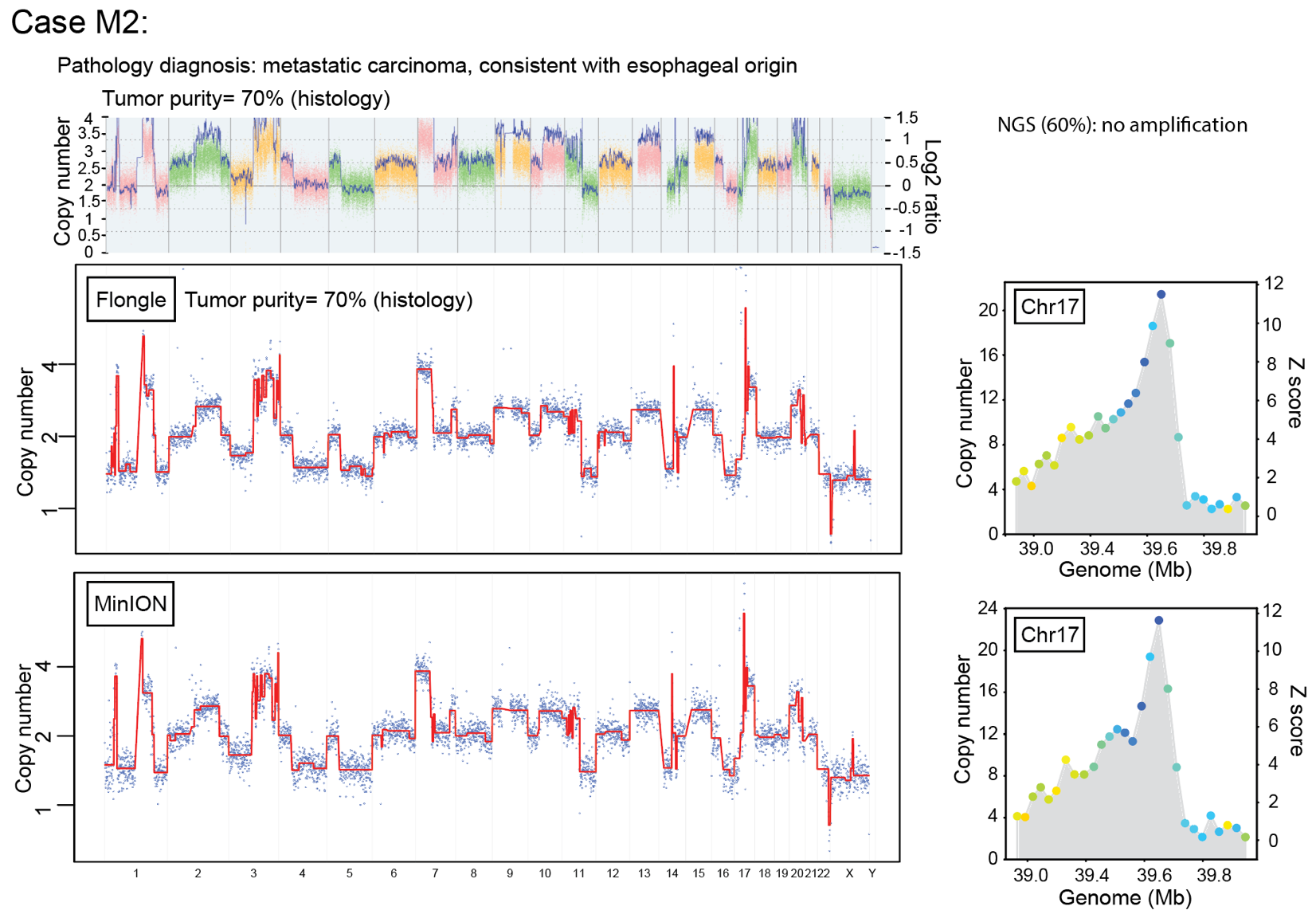

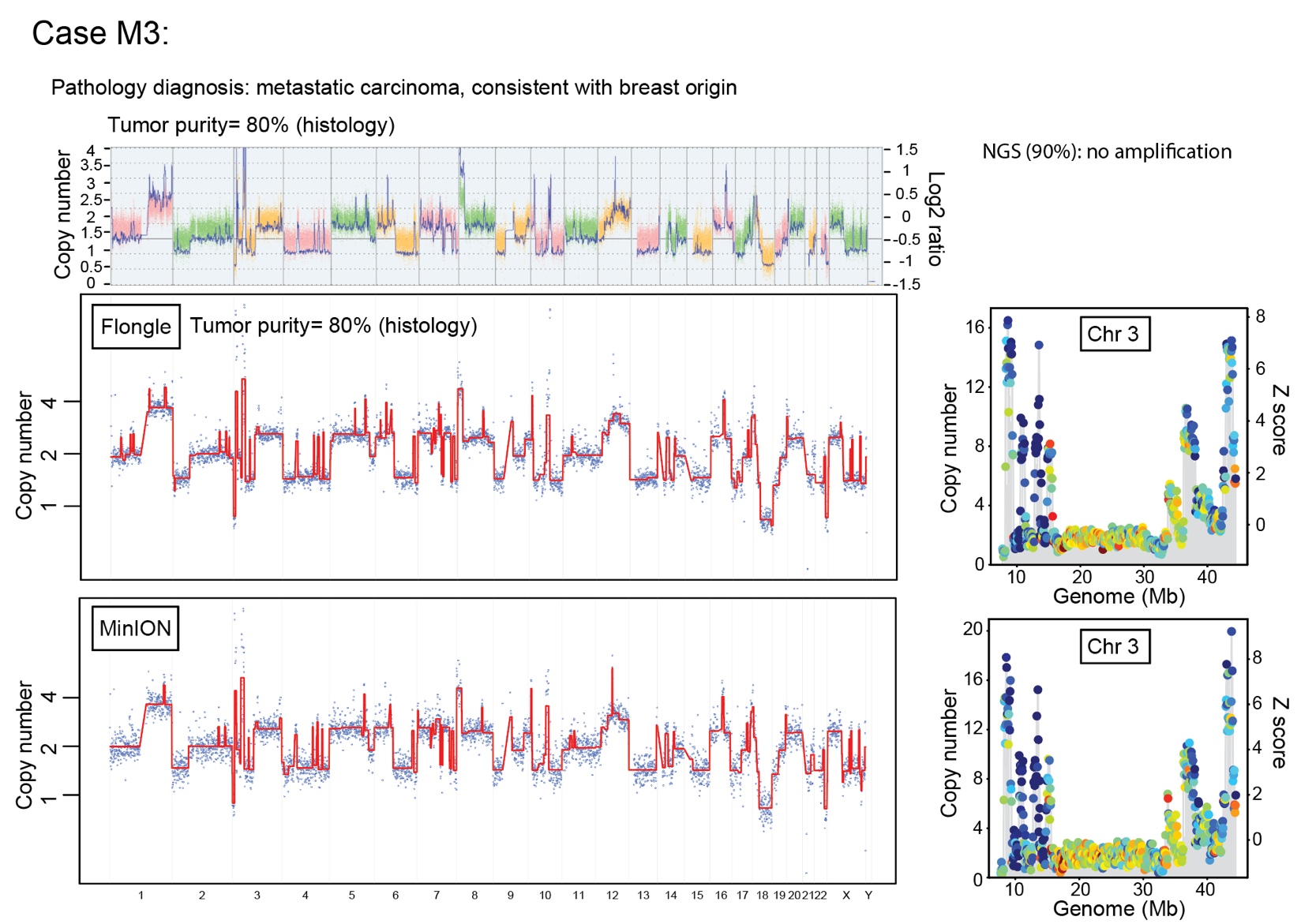

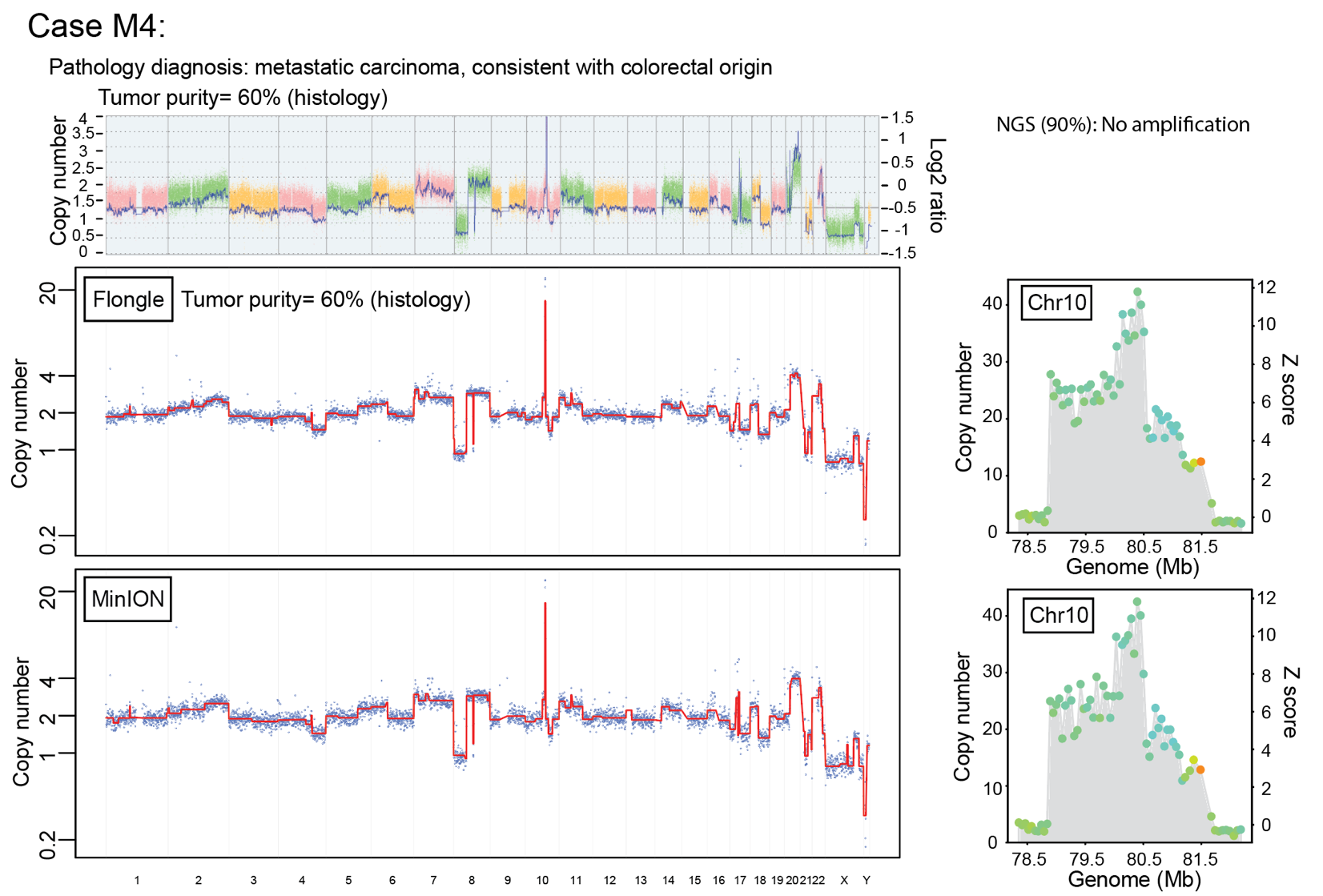

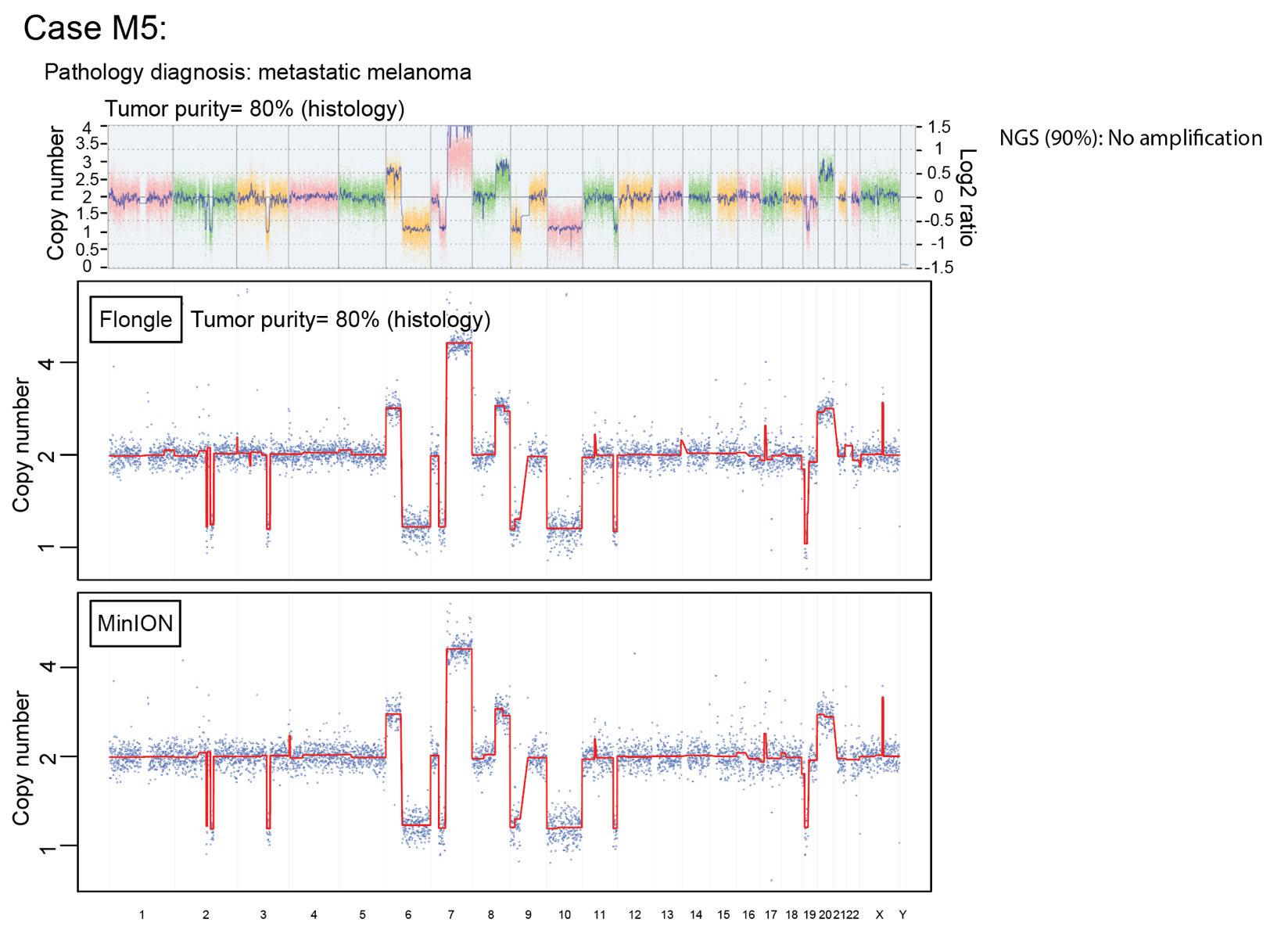

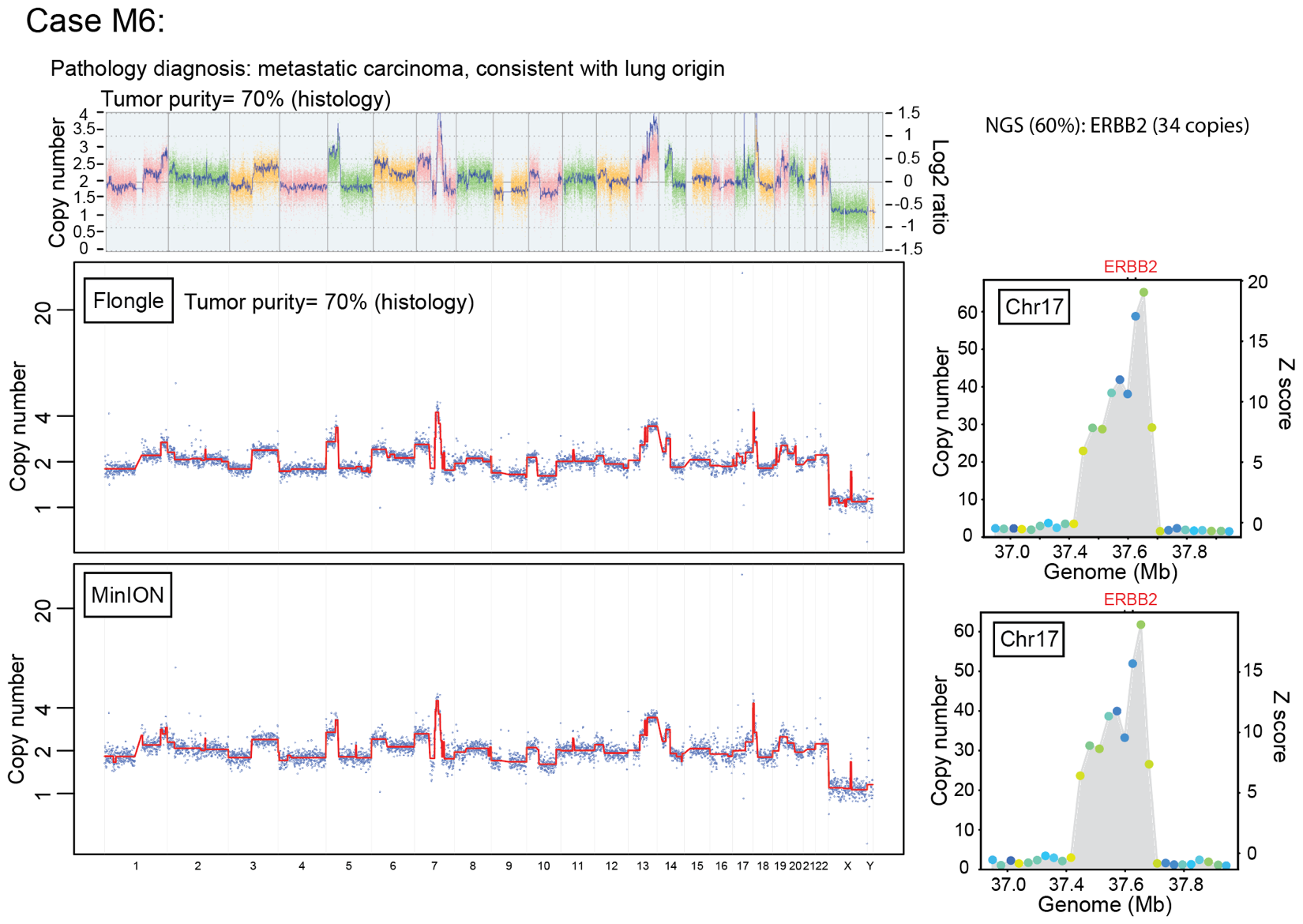

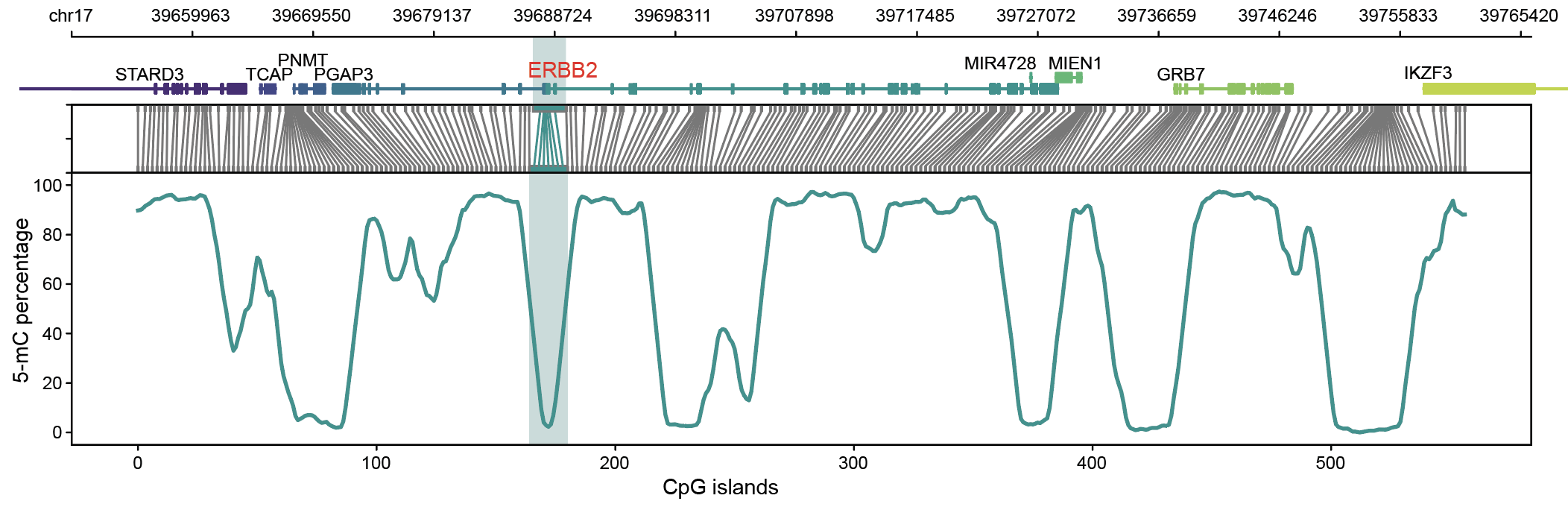

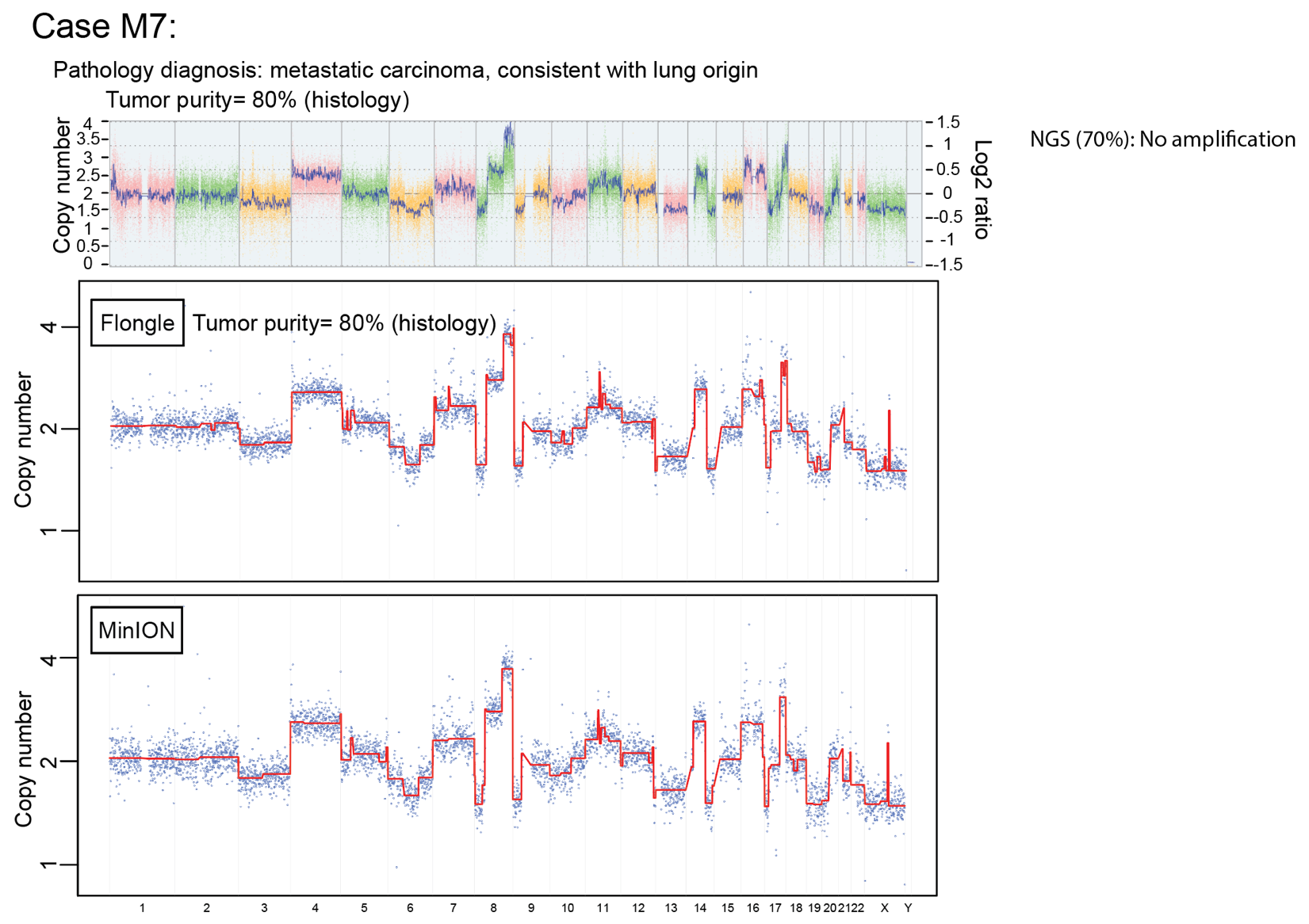

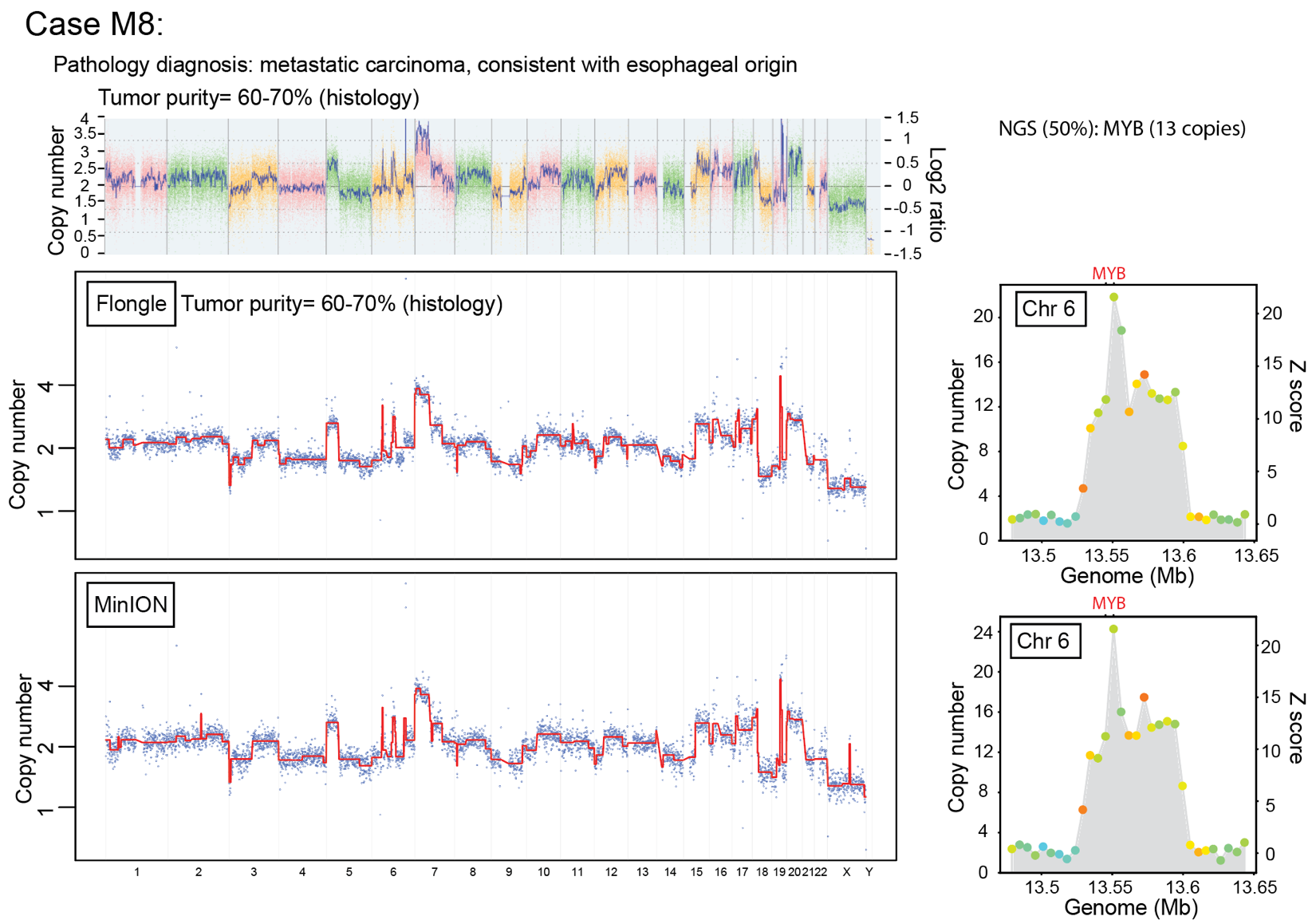

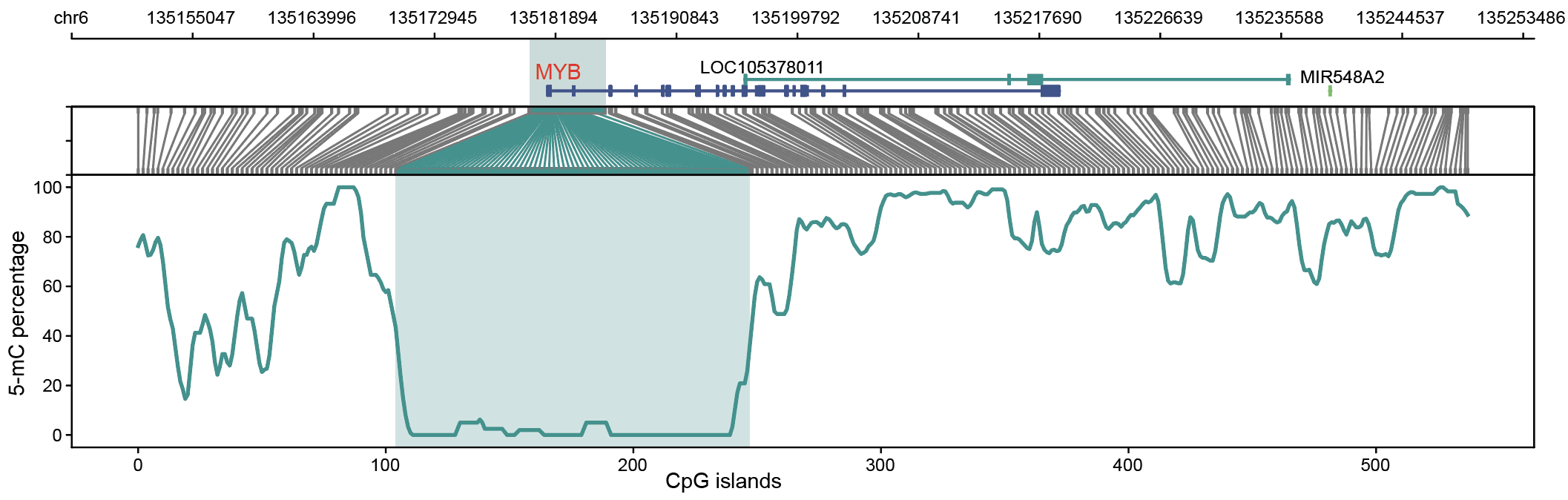

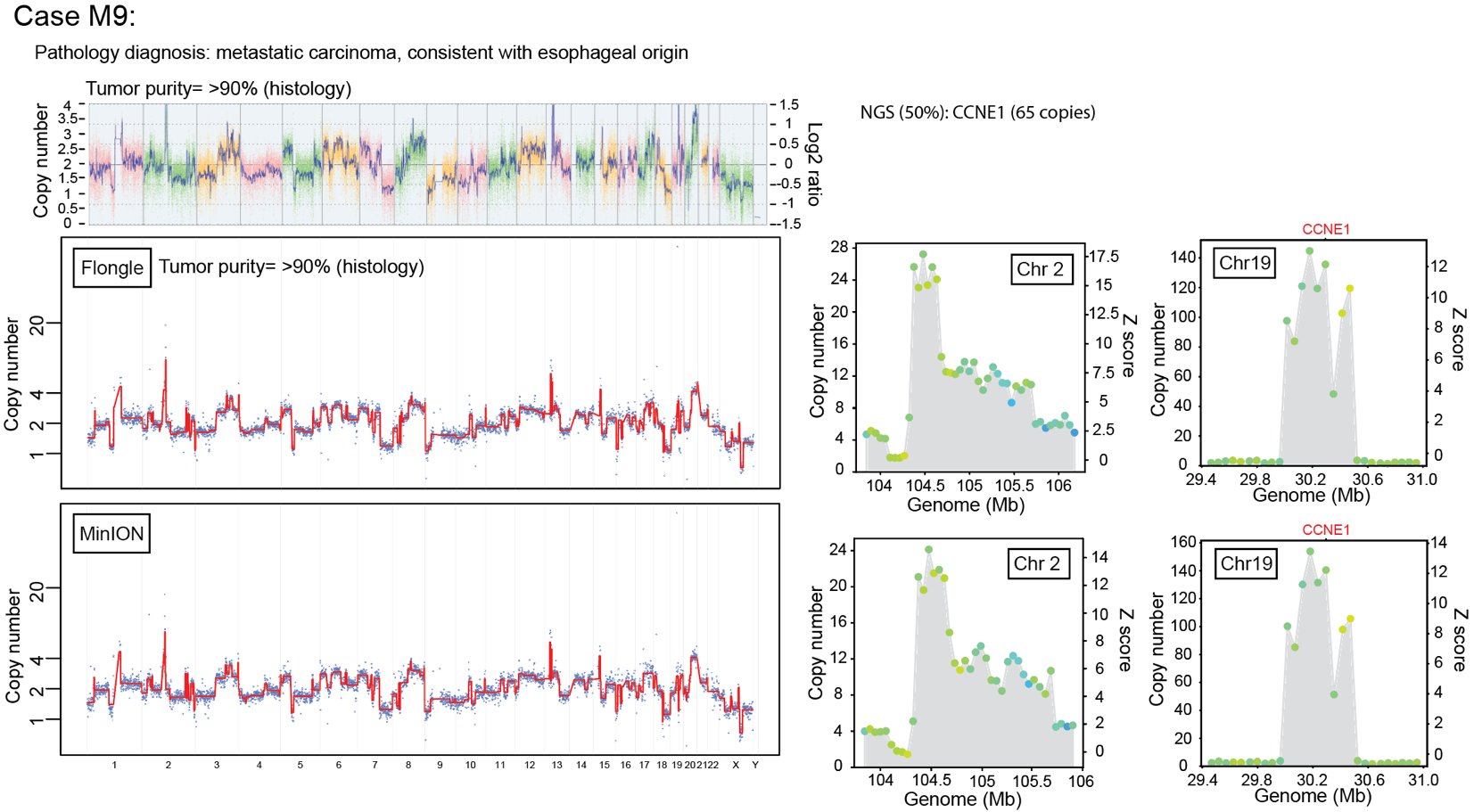

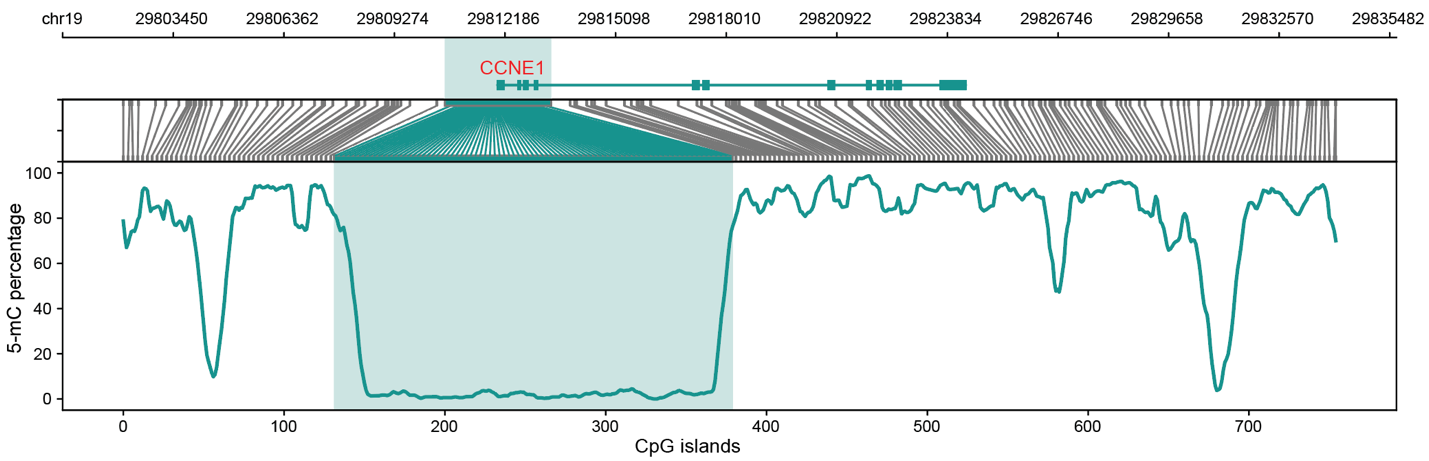
